# Context-specific emergence and growth of the SARS-CoV-2 Delta variant

**DOI:** 10.1101/2021.12.14.21267606

**Authors:** John T. McCrone, Verity Hill, Sumali Bajaj, Rosario Evans Pena, Ben C. Lambert, Rhys Inward, Samir Bhatt, Erik Volz, Christopher Ruis, Simon Dellicour, Guy Baele, Alexander E. Zarebski, Adam Sadilek, Neo Wu, Aaron Schneider, Xiang Ji, Jayna Raghwani, Ben Jackson, Rachel Colquhoun, Áine O’Toole, Thomas P. Peacock, Kate Twohig, Simon Thelwall, Gavin Dabrera, Richard Myers, The COVID-19 genomics UK (COG-UK) consortium, Nuno R. Faria, Carmen Huber, Isaac I. Bogoch, Kamran Khan, Louis du Plessis, Jeffrey C. Barrett, David M. Aanensen, Wendy S. Barclay, Meera Chand, Thomas Connor, Nicholas J. Loman, Marc A. Suchard, Oliver G. Pybus, Andrew Rambaut, Moritz U.G. Kraemer

## Abstract

The Delta variant of concern of SARS-CoV-2 has spread globally causing large outbreaks and resurgences of COVID-19 cases^1–3^. The emergence of Delta in the UK occurred on the background of a heterogeneous landscape of immunity and relaxation of non-pharmaceutical interventions^4,5^. Here we analyse 52,992 Delta genomes from England in combination with 93,649 global genomes to reconstruct the emergence of Delta, and quantify its introduction to and regional dissemination across England, in the context of changing travel and social restrictions. Through analysis of human movement, contact tracing, and virus genomic data, we find that the focus of geographic expansion of Delta shifted from India to a more global pattern in early May 2021. In England, Delta lineages were introduced >1,000 times and spread nationally as non-pharmaceutical interventions were relaxed. We find that hotel quarantine for travellers from India reduced onward transmission from importations; however the transmission chains that later dominated the Delta wave in England had been already seeded before restrictions were introduced. In England, increasing inter-regional travel drove Delta’s nationwide dissemination, with some cities receiving >2,000 observable lineage introductions from other regions. Subsequently, increased levels of local population mixing, not the number of importations, was associated with faster relative growth of Delta. Among US states, we find that regions that previously experienced large waves also had faster Delta growth rates, and a model including interactions between immunity and human behaviour could accurately predict the rise of Delta there. Delta’s invasion dynamics depended on fine scale spatial heterogeneity in immunity and contact patterns and our findings will inform optimal spatial interventions to reduce transmission of current and future VOCs such as Omicron.

## Main text

The SARS-CoV-2 pandemic has been characterized by the appearance and spread of genetically distinct variants that are often associated with faster growth than pre-existing lineages. In May 2021, the World Health Organisation (WHO) announced a new Variant of Concern (VOC), named Delta (Pango lineage B.1.617.2*). Retrospective investigation revealed that Delta was first detected in India in mid-September 2020; it subsequently became the variant primarily responsible for a wave of transmission and mortality in India in early-mid 2021, replacing Alpha and Kappa in the process^6,7^. Reports indicate that Delta has increased transmissibility^8–10^, rates of hospitalisation^11,12^, and immune evasion^13–15^ compared to Alpha (Pango lineage B.1.1.7)^16–21^, the variant previously dominant in many countries. These phenotypes are attributed to a constellation of 30 mutations across the virus genome (Table S1) compared to the reference sequence Wuhan-1, including the spike mutations P681R in the furin cleavage site, thought to increase the speed and efficiency with which the virus fuses with host cells^22,23^, L452R in the receptor-binding domain (RBD), thought to reduce antibody neutralisation^24^, and the nucleocapsid mutation R203M, thought to increase virion infectivity^25^. Delta rapidly disseminated from India to locations worldwide and has been detected in 132 countries, as of September 15, 2021^26^. Delta became the dominant lineage in the UK by mid May 2021^8^, and similar increases in frequency have been observed in other countries worldwide (e.g. ^27,28^).

The emergence of Delta in the UK occurred in the context of a heterogeneous landscape of prior immunity (from infection and vaccination), and non-pharmaceutical interventions (NPIs). Here we examine virus genomes generated from random samples collected during community-based COVID-19 testing in England, between March 12 and June 15, 2021. Our data include 52,992 Delta VOC genomes from England with known dates and locations of sampling, representing >40% of all positive tests in England during the study period (lateral flow and PCR tests; see Methods and details in ^29^). Using these data we evaluate the effectiveness of policies in reducing international importations and how they contributed to the establishment and local transmission dynamics of Delta in England. We then investigate, at a high spatial resolution, how immunity and human mobility contributed to context-specific growth of Delta in England and the United States.

### Reconstruction of international importation and national spread of Delta in England

To provide global context for the emergence of Delta in the UK, we first conducted a phylodynamic analysis of 975 Delta SARS-CoV-2 genome sequences sampled evenly by collection date between March 4, 2021 and June 15, 2021. Details of the origin and spread of Delta within India are still uncertain but coincided with a substantial increase in genomic surveillance across the country which will likely facilitate the study of these important events, but is outside the scope of this work. However, to put the UK epidemic into context we estimate the time of the most common recent ancestor (TMRCA) of Delta globally to be October 19, 2020 (95% highest posterior density [HPD] interval: 2020-09-06 - 2020-11-29). The frequency of Delta in India does not appear to increase substantially until March 2021 (Fig. 1), coinciding with a rapid expansion in case numbers there (Extended Data Fig. 1) and a decline in the relative frequency of genomes assigned to B.1.617.1 (Kappa, a sibling lineage of Delta)^1^. Genomic surveillance in India revealed that several sub-lineages of Delta existed prior to its expansion in March (Fig. 1a,b)^5^. This standing diversity is consistent with undetected transmission of Delta in India between late 2020 and March 2021.

**Figure 1:**
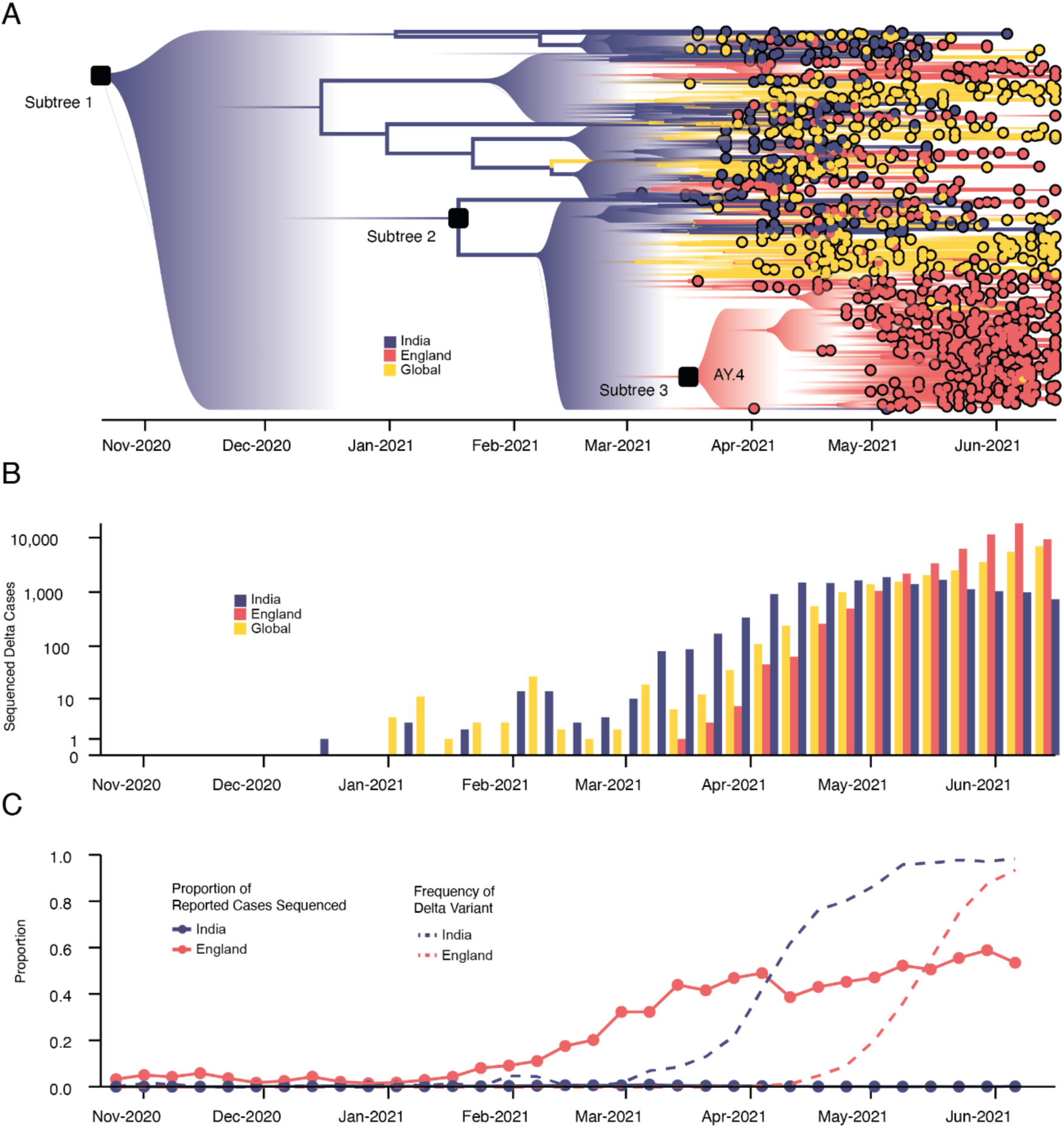
Emergence and rapid geographic expansion of Delta: a) Time-calibrated phylogenetic reconstruction of Delta based on 1,000 sequences subsampled from 93,649 sequences from 100 countries (52,992 from England). The tree was split in 3 subtrees (n=28,783, 28,715, and 36,151 sequences) prior to full analysis. The roots of these 3 subtrees, and of lineage AY.4 are labeled. Lineage colors represent the inferred countries and/or regions where transmission occurred. b) Number of sequenced cases of Delta per week in India, England, and the rest of the world. c) Time-varying proportion of sequenced reported positive cases in India and England (solid lines, n = 52,992 sequences are from England, corresponding to 84% of all sequences from the UK) and the proportion of sequenced cases classified as Delta in India and England (dashed lines).

We evaluated the global dissemination of Delta from March 2021 by multiplying, for each country, estimated numbers of SARS-CoV-2 cases, relative frequencies of Delta, and relative numbers of outward international passengers (Estimated Exportation Intensity, EEI, see Methods). The EEI of Delta climbed rapidly during March and was highest around late April, coinciding with peak incidence in India (Extended Data Fig. 2). Subsequent rapid growth of Delta in the USA, Russia, UK and Mexico, and its decline in India resulted in the former locations becoming the main exporters of Delta by June 2021 (Extended Data Fig. 2), corroborating global trends in Delta phylogeography (Fig 1a) and reported cases (Fig 1b). Similar patterns of rapidly changing foci of international dissemination were observed for the initial wave of SARS-CoV-2 in 2020^30,31^.

To evaluate the temporal dynamics of Delta importation into England and to reconstruct its subsequent local spread, we conducted a travel history-aware Bayesian phylogeographic analysis^32^ of 93,649 Delta sequences, from GISAID and COG-UK, which accounts in part for the phylogenetic uncertainty inherent in SARS-CoV-2 phylogenies^31^. To render the analysis tractable we split the full tree into three independent subtrees (Fig. 1a) prior to phylogeographic analysis. Virus genomes were generated from ∼40-60% of all positive cases in England during the emergence of Delta between March and May 2021 (Fig. 1c)^33^, providing a unique opportunity to characterize the virus’ spread at a high spatio-temporal resolution^33^.

We estimate a minimum of 1,458 (95% HPD: 1398-1513) separate international introductions of Delta into England, with approximately half inferred to have originated from India (posterior mean 56.5% ; 95% HPD 53.7%-59.1%). We find the majority of English Delta genomes can be traced back to introductions that are inferred to have occurred prior to the implementation of a mandatory hotel quarantine for people arriving from India (posterior mean 84.3%; 95% HPD: 77.8-90.4%). During this period 90.0% of introductions are inferred to have originated from India (95% HPD: 86.5-93.1%). These inferred importation dynamics closely follow individual-level travel histories from infected incoming international passengers (Fig. 2b).

**Figure 2:**
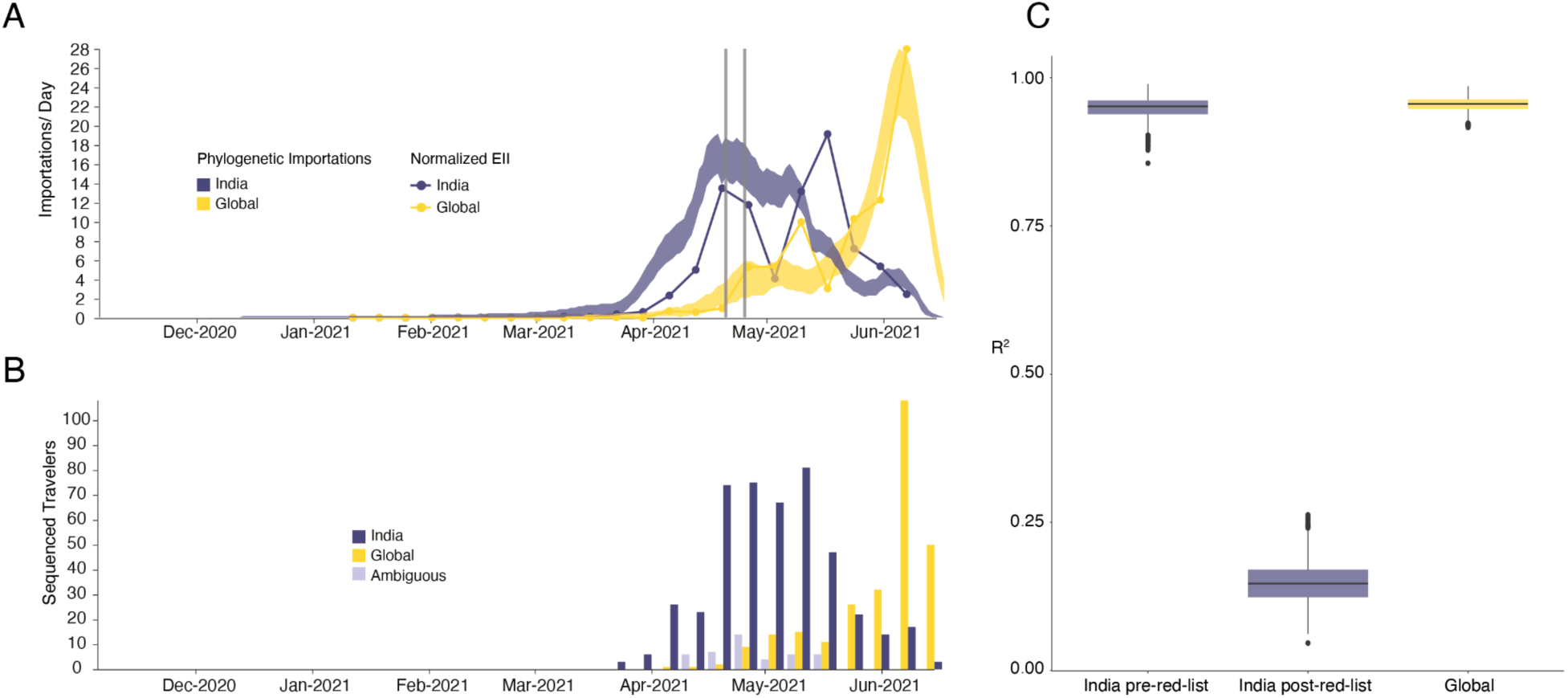
Timing of importations of Delta into England. **a)** Daily number of estimated importations of Delta from India (blue shaded area) and other countries (yellow shaded area) inferred from phylogenetic analysis. Shaded areas show 95% HPDs of the estimate. Blue and yellow lines show the Estimated Importation Intensity (EII) of Delta, obtained by combining data on human movements, cases, and prevalence of Delta, normalized to the same scale as the phylogenetic estimates. Grey vertical lines indicate the announcement of travel restrictions from India to England (April 18, 2021) and its implementation on April 23, 2021. **b)** Temporal distribution of genome sequences from cases with known travel history from India (blue) and other countries (yellow). Isolates with recent travel to both India and other countries are considered ambiguous (lavender) **c)** R^2^ (coefficient of determination) between estimated number of importations from the phylogenetic analysis and the Estimated Importation Intensity (EII) (panel a). The R^2^ is calculated separately for India (blue) before and after hotel quarantine was introduced, and for other countries (yellow), whilst also accounting for phylogenetic uncertainty.

High variation in sampling intensity among countries means the true number of importations into England is likely much larger than that inferred from phylogeographic analysis alone (Fig. 1b & c, see related discussion in the context of the UK’s first wave^31^). For example, the AY.4 lineage (Fig 1a) comprises 42,445 sequences and was likely imported to England many times. We investigated AY.4 by pairing genomic data with contact tracing data collated by Public Health England. During the study period we found 61 AY.4 sequenced cases had a travel history from India and 140 had a travel history from elsewhere; similar to the time-varying importation dynamics seen across the entire dataset (Fig. 2a; Extended Data Figure 3). Hence sampling heterogeneity means that the number of importations estimated from phylogenetic analysis represents a lower bound on the true number^31^.

To investigate the importation of Delta into England specifically, and to cross-validate the results above using independent data, we estimate the Estimated Importation Intensity (EII) of Delta to England through time^31,34^. The EII is a metric of Delta importation that represents trends in the number of Delta cases arriving in the country, irrespective of whether or not those cases result in local transmission. Contrastingly, the phylogenetic analysis above better captures trends in the number of Delta introductions that did lead to forward transmission in England. The EII combines (i) weekly reported cases, (ii) weekly prevalence of Delta genomes, and (iii) weekly aggregate human mobility (inferred from mobile phones) into England via direct connections (Fig. 2a; see ^31,34^ for related approaches). The EII from India increased rapidly in April 2021 following the rise in cases in India and remained high until the end of May 2021. However, we observe that the correlation between the EII and the numbers of importations inferred from phylogenetic analysis declined significantly after the implementation of hotel quarantine for travellers from India (Fig. 2c, mean R^2,before^ = 0.95 and R^2,after^ = 0.15), indicating that this intervention reduced the number transmission chains established locally per infected incoming traveller. From late May importations from countries other than India dominated Delta importation into England (Fig. 2a), a trend also visible in contact tracing data carried out by Public Health England (Fig. 2b, R^2^ between non-India importations and EII = 0.95, Fig. 2c). Even though we observe that the implementation of hotel quarantine was effective in reducing onward transmission, substantial importation had already occurred before its implementation and additional introductions from other countries likely further accelerated the spread of Delta in England from May onwards.

There are several reasons why some importations led to onward transmission within England after the implementation of hotel quarantine for arriving travellers: (i) a separate terminal for arrivals from mandatory quarantine countries was not opened at the UK’s largest airport (London Heathrow) until 1st June^35^, so arriving passengers may have mixed with others prior to initiating mandatory quarantine; (ii) individuals may have become infectious and transmitted only after leaving quarantine, either due to an unusually long latent period or within-group transmission during the quarantine period, although we do not consider this probable^36^; (iii) individuals may infect others on a connecting flight where the connecting airport did not require hotel quarantine; (iv) there were exemptions to hotel quarantine that may have led to onward transmission in the community^36^.

### Transmission lineage dynamics, dissemination, and establishment of Delta in England

Importations of Delta occurred on a background of relaxation of social distancing in England: on April 12th outdoor dining and non-essential retail reopened, and on May 17th restrictions on indoor dining and international travel were relaxed^37^. The relative frequency of Delta genomes in England increased rapidly during May and reported COVID-19 cases subsequently increased^38^ (Fig. 1c). Initially, Delta transmission clusters were concentrated in the North West of England and were commonly associated with returning travellers^17,39,40^. We sought to reconstruct the internal dispersal dynamics of independently-imported Delta transmission lineages in England, in the context of changing non-pharmaceutical interventions.

We analysed all identified Delta transmission lineages in England using continuous phylogeography, thereby inferring their history of dissemination among subnational regions (UTLAs; upper tier local authorities). We observe high heterogeneity among ULTAs in the numbers of Delta introductions from other English regions (Fig. 3a), with Lancashire and Greater Manchester each receiving >2000 estimated independent introductions and Torbay only 9. The majority (n = 11,960) of Delta sequences in England belong to a single transmission lineage (lineage I, Fig. 3d), which was sampled mostly in Greater Manchester and Lancashire, and we observe many short-range lineage movements among UTLAs in these areas (Fig. 3a). Greater London also received many Delta cases from elsewhere in England (Fig. 3a), as expected, given its population size and connectedness to other metropolitan areas^34^. Transmission lineages II and III each comprise 3000-4000 genomes; the former is distributed across multiple urban areas (especially in the North West) whilst the latter is focussed in Greater London and the South East (Fig. 3d). We also highlight transmission lineage V (Fig. 3d), originally centered in Bedfordshire, the location of one of the first Delta outbreaks in England and was subjected to surge testing ^41^ (Extended Data Fig. 5).

**Figure 3:**
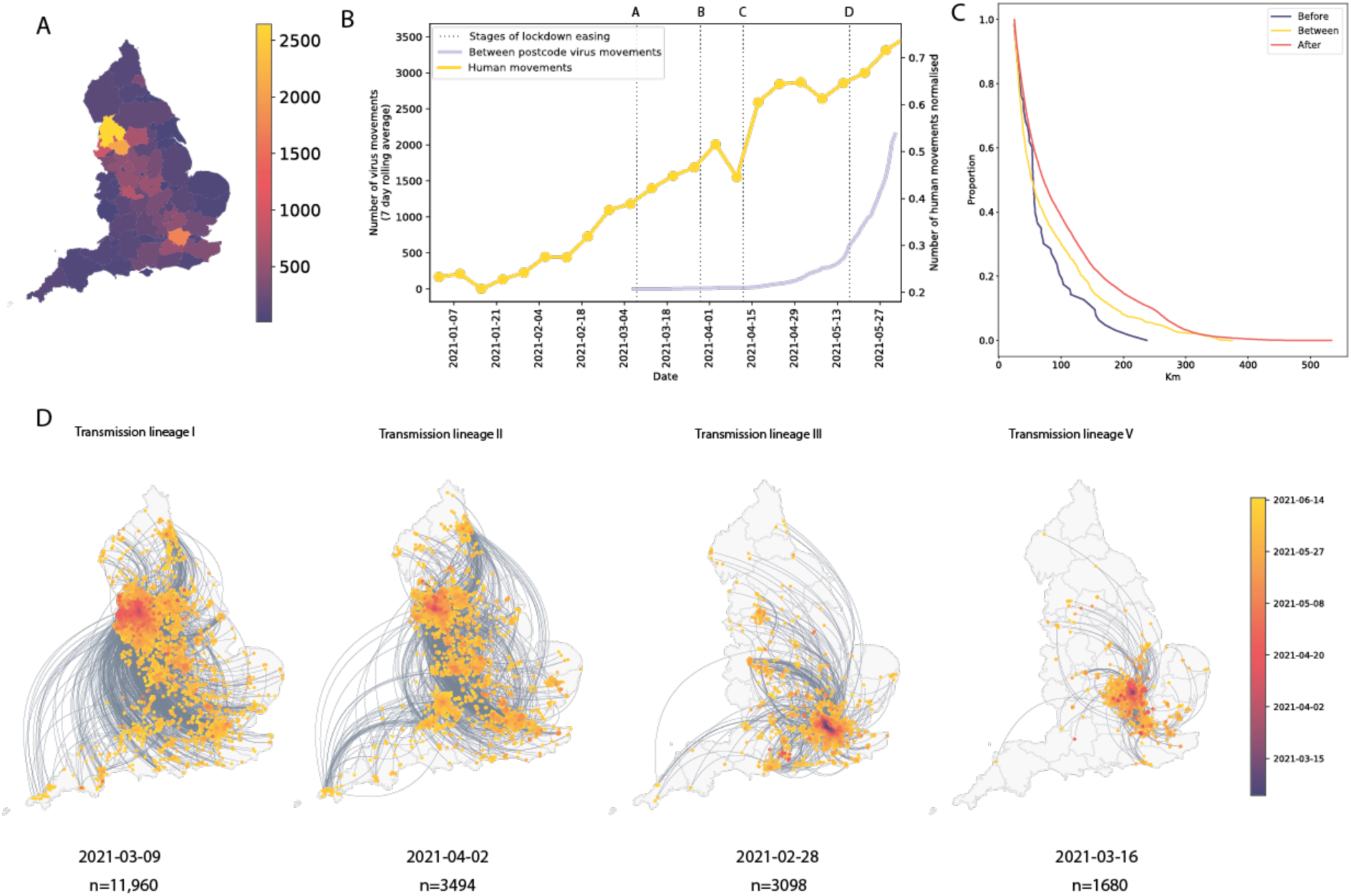
Introductions and regional dynamics of Delta transmission lineages. **a)** Number of independent introductions per UTLA in England based on continuous phylogeographic analysis of all Delta transmission lineages with >5 sequences. **b)** Trends in aggregate human mobility and the number of virus lineage movements among postcode districts. Letters denote stages of lockdown easing: A (2021-03-08) schools reopen and limited mixing between households outdoors permitted; B (2021-03-29) “Stay at home” directive lifted, more outdoor mixing allowed (up to six people from two households; C (2021-04-12) non-essential retail re-opened, outdoor dining permitted, holiday lets and campsites re-open; D (2021-05-17) indoor hospitality opens, indoor mixing permitted. **c)** Proportion of virus lineage movements between postcodes >25 km apart: y-axis denotes the proportion of movements that are less than or equal to the value on the x-axis. This is shown for movements before lockdown easing on 12th April (blue), between 12th April and 17th May (yellow) and after 17th May (red). **d)** Virus lineage movements inferred by continuous phylogeographic analysis for four large transmission lineages (see transmission lineages IV-VII in Extended Data Fig. 5). Direction of lineage movement is anti-clockwise, and dots represent the start and end points of movement, coloured by inferred date. The size and inferred TMRCA of each lineage is shown below each map. Distance kernels for each lineage can be found in Extended Data Fig. 7.

In early May, the number of virus lineage movements among locations accelerated (Fig. 3b, Extended Data Fig. 6), showing that growth in Delta frequency (Fig. 1c) was associated with regional dissemination. This spread occurred on the background of relaxing NPIs and increased mixing (between mid-January and June 2021, mobility in England increased from 20% to 70% of its pre-pandemic level and estimated mean daily contacts rose from ∼2 to ∼5, ^42^). In contrast, the initial wave of SARS-CoV-2 introductions to the UK, in spring 2020, occurred during a period of increasing travel and social restrictions^31^. In general we find that, as NPIs were progressively relaxed through time, long-range viral lineage movements comprised an increasing proportion of all movements (Fig. 3c).

For the seven largest Delta transmission lineages in England (I-VII) we observed ∼3 times more exports from Greater Manchester than from Greater London. This difference matches early epidemiological data: the largest and earliest Delta outbreaks were located in the North West (on May 21 Bolton had 452 cases per 100,000 whilst Greater London had 21.6 ^43^, see Methods). Introductions of Delta into other, smaller urban areas also spread rapidly (e.g. transmission lineage V, Fig. 3d) and were important for the propagation of the variant across England. We observe spatial structure of the seven largest lineages where the frequency of viral movements decline by distance away from the origin location but we also observe a second peak at ∼260km (similar to the distance between Greater London and Greater Manchester, Extended Data Fig. 7). Although North West England was a focus of early Delta transmission, the Delta epidemic in England derived from many successful independent international importations. Each of the main Delta transmission lineages in England grew at a similar rate (Extended Data Fig. 4). In contrast, the Alpha variant (Pango lineage B.1.1.7) expanded across the UK from a single origin in South East England^34^. The spatial expansion of Delta transmission lineages plateaued after early June, when most UTLAs had established Delta transmission and the relative frequency of Delta genomes in England had exceeded 90%^44^.

Although Scotland, Wales or Northern Ireland are not included here, case count data suggests that cities in England^45^ were the main source of the expanding Delta epidemic in the UK; due to this source-sink structure we do not anticipate that omitting these countries substantially affects our reconstruction of epidemic dynamics in England (of the Delta genomes available before 15th June 2021, 57,592 were from England, 9738 from Scotland, 1067 from Wales and 325 from Northern Ireland).

### Investigating the factors contributing to accelerated growth of Delta

Regional and international heterogeneity in incidence, vaccination, and human mobility have been shown to determine the dynamics of infectious diseases^46^, including those of SARS-CoV-2^31,47–52^. We use a combination of epidemiological, aggregate human mobility, and genomic data to test the hypothesis that (i) relaxation of NPIs impacted Delta local growth rates in England, and (ii) immunity from infection and vaccination affected Delta growth in the US. To do so we develop a hierarchical Bayesian model to estimate the impact of these factors on the weekly relative growth of Delta (i.e., the weekly change in the observed proportion of Delta genomes on a log odds scale)^53^ at the UTLA level for England and the state level for the US. Models for estimating the increase in transmissibility of new variants are typically based on increases in relative frequency^3,16,53–55^ but rarely take into account other potential confounding factors, such as population immunity^56^.

In general, growth rates varied widely across locations and weeks in England (Fig. 4). This variation may be explained in some cases by specific events, such as the beginning of university holidays in May and June 2021 (e.g. Oxfordshire, Fig. 4a, b). Our model estimates that the most important tested predictor of the variation in growth of Delta (relative to Alpha) across UTLAs in England was within-UTLA mixing (i.e., relative changes in weekly within-UTLA human mobility compared to the pre-pandemic period, Figure 4a, Table S4). The importance of this factor is unsurprising, as preemptive restrictions on movement and social mixing slow the emergence of new pathogens or variants^57^ (see counterfactual scenarios in Extended Data Fig. 9); the cost/benefit ratio of such restrictions will of course depend on the specific context of variant emergence. The relaxation of NPIs therefore increased both within- and among-region transmission (see Fig. 3c). Other European countries did not observe such a rapid increase in Delta relative frequency during May 2021^1^; possible reasons for this difference are (i) during that time levels of mobility and mixing (both local and regional) were lower in those countries and/or (ii) those countries potentially received fewer international importations of Delta (86,489 passengers flew from India to the UK between March and June, whilst 43,515 flew to Germany, and 16,688 to France, during the same time). Vaccination rates did not explain local variation in Delta growth rates in England, possibly because there was insufficient heterogeneity in vaccination rates among UTLAs to detect any effect^58^.

**Figure 4:**
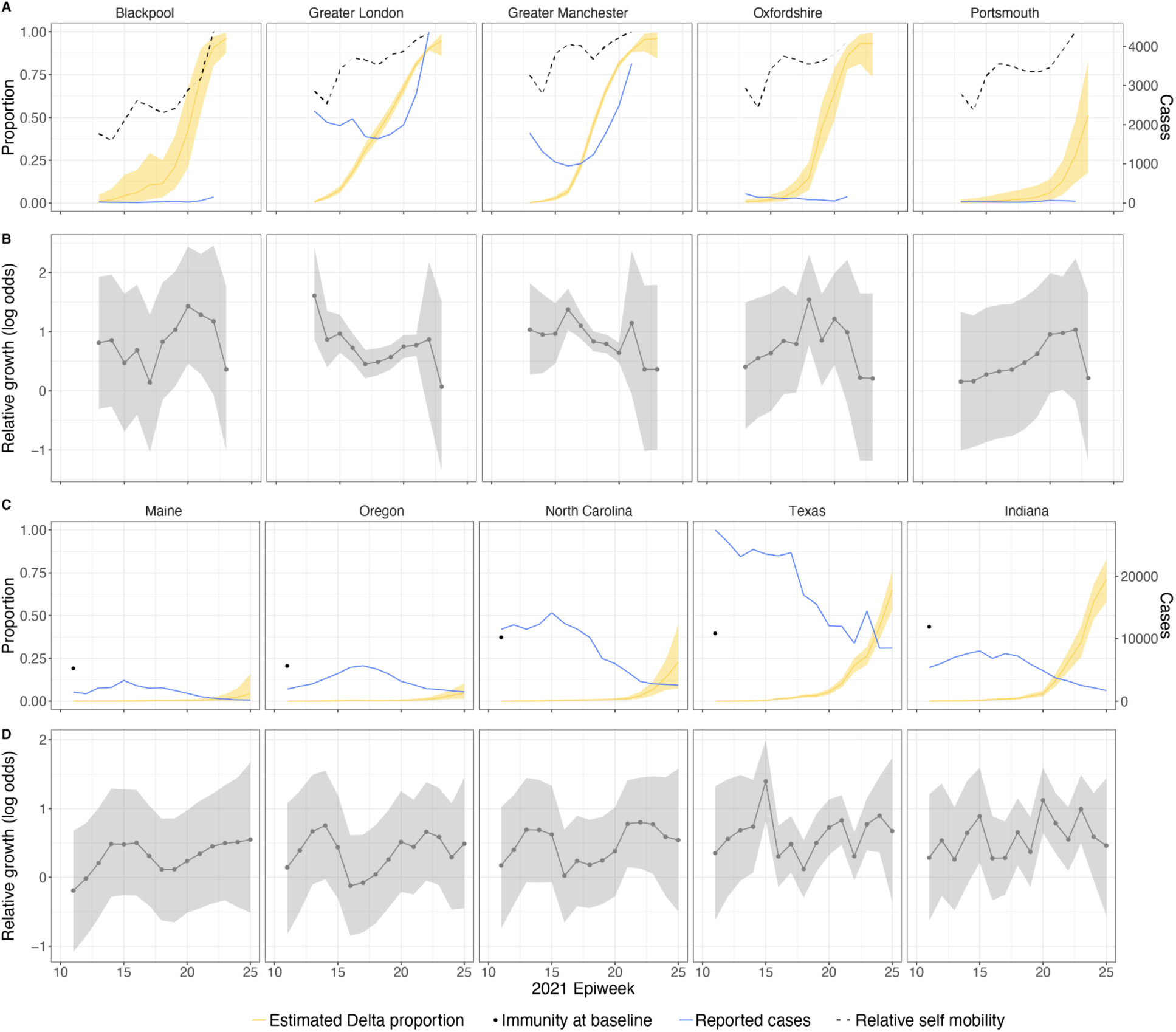
Delta growth rates among regions in England and USA. a) Estimated proportion of genomes that belong to the Delta variant (yellow) for several illustrative UTLAs in England, and observed relative aggregate human mobility (black dashed line), number of reported cases (blue line). **b)** Corresponding time-varying relative growth of Delta (on the log odds scale). The light shaded regions in all plots represent 95% Bayesian credible intervals. **c)** Estimated proportion of Delta variant samples across several illustrative US states (yellow), reported number of cases (blue line) and observed immunity at baseline (black dot). **d)** Corresponding time-varying relative growth of Delta (on the log odds scale).

Among US states, levels of immunity through infection and vaccination (as measured by fraction of people with two vaccine doses) varied considerably (from 15.2% at baseline, 14th March 2021, to 56.4% at week ending 26th June 2021^59^). Using our model while accounting for local mixing patterns, we find that higher baseline local immunity levels were associated with higher overall growth of Delta relative to other lineages (Fig. 4c, d, Table S4). This observation is superficially counter-intuitive but has several possible explanations: (i) due to social and demographic variation, pathogens can exhibit different R_0_ values in different locations, hence locations with high levels of previous exposure are more likely to support faster transmission of a newly introduced VOCs (provided that sufficient numbers of local susceptibles remain); (ii) Delta is better able to evade neutralising antibodies than other co-circulating variants, specifically Alpha^5,60^. Whilst this hypothesis cannot be excluded, it cannot explain the replacement of Beta by Delta in South Africa^61^ and Delta’s success is better explained by its increased intrinsic transmissibility than by its ability to evade immunity^5,10,62–64;^ (iii) aggregating data to the US state level may obscure inference of epidemiological dynamics, which may vary substantially at local scales due to variation in vaccination or behaviour^65,66^. In a sensitivity analysis (Table S5) we consider only immunity from prior exposure^67^ (not vaccination) and find similar trends. The magnitude of the effect of prior immunity and human mobility can be seen in counterfactual scenarios in Extended Data Figs. 9 and 12.

Using model comparison and out-of-sample prediction (withholding data from the final few weeks), we find that models that included predictors such as baseline immunity and vaccination (US) and within-UTLA mobility (England) fit the observed trajectory of Delta relative growth better than a model without covariates (Methods and validation, Supplementary Information, Extended Data Figs. 10, 11, 13 & 14, Extended Data Tables 6 & 7). We refrained from translating estimates of the growth rate of Delta relative frequency into differences in the reproduction number, as this is sensitive to assumptions about the generation time of the variant, which is also influenced by NPIs and immunity^68^. At the time of analysis, there was no consensus on the generation time of Delta. Further studies should consider estimating the generation times of VOCs in specific contexts of immunity, NPIs and household structure to accurately translate relative growth rates into R_t_^69^.

### Discussion, limitations and future work

We find that growing epidemics of SARS-CoV-2 Delta worldwide led to a wave of importations of the VOC into England, initially from India, and later from other countries. These importations found fertile ground as they arrived in a context of easing social restrictions, and consequently expanded rapidly across England. Much transmission occurred in unvaccinated and younger populations^70^, and high levels of Delta transmission within the UK led to onward dissemination of the variant to other countries (e.g. ^71^). By pairing the phylogenetic results with contact tracing data we conclude that hotel quarantine measures were effective in reducing onward transmission of imported Delta cases in England. However, after May 21, we found that levels of local social mixing in England, not the number of importations, was associated with faster relative growth of Delta. At that point the independently introduced transmission lineages grew at a similar pace; details of their geographic distribution and expansion will support future work defining the optimal spatial interventions to reduce transmission of VOCs in England.

Undetected genetic diversity and uneven sampling of Delta in India make precise estimation of the number of importations to England difficult from genetic data alone^72^ (Extended Data Fig. 8). However, our phylogenetic estimates strongly correlate with estimates derived from independent data on case incidence, Delta prevalence, and arriving travellers (EII, Methods, Fig. 2c) during the period before quarantine policies were announced. Fortunately, additional contact tracing data from public health agencies allowed us to overcome limitations inherent in the unevenly sampled global virus genomic data set, and provide additional confidence in our findings.

Our statistical analysis shows that higher Delta growth rates were positively associated with levels of population immunity and vaccination in the United States and with levels of local mixing in England. In the future, the existence or magnitude of NPIs needed to reduce the healthcare burden of Delta (or future VOCs) to sustainable levels will depend on local levels of population immunity (through vaccination and prior infection). Future work should focus on which factors are most conducive to spread in particular contexts (e.g., high vs. low NPI regimes and across levels of population immunity) so that responses can be planned accordingly. This requires a better characterisation of the distribution and variation of infectiousness through time, and an understanding of virus generation time in different behavioural contexts^73^, for example amongst individuals who are vaccinated, unvaccinated and/or had previous exposure to SARS-CoV-2 (including with which lineage). To do so effectively will require investments in large-scale and coordinated serological studies^74^ especially for VOCs with ability to evade immunity.

Even though global reporting of case numbers, virus genomic surveillance, sampling strategies and mobile phone penetration differ across the world, our estimates can still provide qualitative insights into the trends in the source locations and rates of international importation. Including estimates of likely importations in disease surveillance programmes may help support public health decision making^75^ and further improvements on these estimates can be achieved when global health surveillance systems are more integrated, and investments in data generation and capacity are linked directly into paired genomic-epidemiological analytical pipelines^76^.

The detail with which we document the spatial invasion process of Delta in England provides an opportunity to re-examine how more spatially targeted interventions can support COVID-19 control in the future. Globally coordinated data and analytical pipelines that capture heterogeneity in virus circulation, immunity and policy responses will be necessary to produce the insights necessary to curb the spread of emerging infectious diseases and new variants. However they can only be successful when integrated into a public health framework that can respond and rapidly adapt to public health threats during their emergence^4,77^.

## Data Availability

UK genome sequences used were generated by the COVID-19 Genomics UK consortium (COG-UK, https://www.cogconsortium.uk/). Data linking COG-IDs to location have been removed to protect privacy, however if you require this data please visit https://www.cogconsortium.uk/contact/ for information on accessing consortium-only data. The Google COVID-19 Aggregated Mobility Research Dataset used for this study is available with permission from Google LLC. Code to reproduce the statistical analyses on Delta growth can be found here: https://github.com/sumalibajaj/Delta-Statistical-analysis-share. The code and accession ids of sequences used to run the phylogenetic analysis as well as an GISAID acknowledgment table are available here: https://github.com/COG-UK/Delta-analysis.

## Methods

### Genomic data

International (non-UK) sequences were downloaded from GISAID on September 15, 2021 and combined with English sequences taken as part of community surveillance (pillar 2) available in COG-UK as of September 2021. Sequences were processed and aligned as part of the daily datapipe analysis managed by CLIMB on behalf of COG-UK. Duplicate and environmental sequences as well as those with impossible or incomplete collection dates were removed. All sequences were aligned to the reference Wuhan-Hu-1 (genbank accession MN908947.3) with minimap2 and samples with less than 93% coverage were discarded. Scorpio (https://github.com/cov-lineages/scorpio) was run as part of Pangolin^78^, and sequences containing the Delta VOC constellation of mutations were kept for further analysis.

Problematic sites in the resulting alignment were masked prior to phylogenetic inference and isolates with known sequence artifacts removed (see https://github.com/COG-UK/Delta-analysis for details). Additionally, mutations in the Delta VOC have caused widespread amplicon drop out of amplicon 72 in the commonly-used ARTIC primer scheme (https://www.protocols.io/view/ncov-2019-sequencing-protocol-v3-locost-bh42j8ye) before the introduction of version 4 of the primer scheme. To avoid spurious phylogenetic associations based on differential treatment of amplicon dropout with COG-UK and across the globe, we masked sites 2142-21990 which represent the region solely covered by amplicon 72 and are not overlapped by neighboring amplicons.

### Phylogenetic analyses

To provide an overview of the global expansion of Delta (Fig. 1a), we analysed a subset of 1,000 Delta genomes sampled evenly through time. To minimize the effect of incorrectly reported collection dates, we restricted our analysis to samples where the lag between sample collection date and GISAID submission date is less than four weeks. To further ensure only the highest quality samples were included, we built an maximum likelihood tree using iqtree2^79^, rooted with Wuhan-Hu-1 (genbank accession MN908947.3) as an outgroup, and used Treetime^80^ to remove tips lying beyond two interquartile ranges from the regression of time against root-to-tip distance. This analysis resulted in a final dataset of 975 samples. The temporal tree estimated by treetime was used as a starting tree in the following Bayesian analysis with slight modifications to randomly resolve polytomies. Two chains of 100 million states were run using BEAST v1.10.4^81^ with sampling every 20,000 states. Both chains were combined with the first 10 million states removed for burnin. We used a HKY+Γ substitution model^82^, a flexible Skygrid coalescent prior^83^ with grid points every two weeks^80^, and an asymmetric, discrete phylogeographic model with samples assigned to Indian, English, and Global locales. Preliminary analysis showed very little temporal signal in the data, which is unsurprising given the relatively slow evolutionary rate of SARS-CoV-2 and the short study period. Therefore, in all analyses the evolutionary rate was fixed to 7.5×10-4 substitutions / site as estimated in^31^. Convergence was assessed using Tracer v1.7^84^.

The goal of our phylogenetic analysis was to accurately and efficiently describe importation dynamics into England, without sacrificing the dense sampling needed to reconstruct internal spread at a high resolution. Due to the large size of the required dataset, we followed a similar phylogenetic approach to that used in^31^. First, an approximately maximum likelihood phylogeny was built using a JC69 substitution model in FastTree^85^, and rooted on Wuhan-Hu-1 (genbank accession MN908947.3), a high quality Pango lineage B sample from 2019-12-26, as an outgroup. Internal branches representing less than one substitution were collapsed to polytomies. This tree was then split into three subtrees of roughly equal size (Fig. 1a) (28,783, 28,715, and 36,151 tips). As above, Treetime^80^ was then used to remove temporal outliers, generate a starting time tree, and estimate the number of mutations along each branch. For subtree an empirical distribution of time trees was estimated independently using a recently implemented model in BEAST v1.10^81^ (commit:d1a45) which replaces the substitution model in classical analyses. Briefly, in this approach the likelihood of the number of mutations along each branch was calculated from a Poisson distribution with mean equal to the evolutionary rate multiplied by the length of the branch in time^86^. In this approach, the standard topological tree search is constrained to operators that sample node heights and resolutions of polytomies present in the substitution tree.

For each subtree, 50 MCMC chains of 40 million iterations were run, sampling trees every 2 million states with the first 20 million states removed as burnin, resulting in datasets of 514-520 empirical trees. The analyses were run using a flexible Skygrid coalescent prior^83^ with grid points every two weeks^80^. Model convergence and proper statistical mixing were verified in Tracer v1.7^84^.

The empirical trees sets estimated above were used to reconstruct importations into England under an asymmetric discrete phylogeographic model. Taxa were split into three locations: England, India and Global, with the Global state representing all countries other than England and India. We used the recently developed travel aware phylogenetic model available in BEASTv1.10^32^ to better inform the transition rates in the reconstructed phylogeography. “Travel history” nodes were placed 1 week before isolates from England with known travel history. Where such travel included both India and other countries, ambiguous non-UK states were used. We ran eight chains of 625,000 states, sampling every 2,250 states and with the first 62,500 states removed as burnin, resulting in a total of 1,998 or 1,999 trees sampled from the posterior distribution.

Introductions were defined as nodes inferred to be in England with parents in either India or the catch-all Global location. The date of importation was assumed to be half-way between such a node and its parent. Five trees in the posterior set were excluded as they placed the root node of subtree 3 in England; this event was deemed highly unlikely as this node lies at least three months prior to the first sample from England during a time at which sequence coverage was above 50% in England. In

Following the importation analysis, the seven largest importations (those with >1500 sequences, n = 25,983) were selected, as well as all importations with five or more sequences, from a representative tree from the posterior set with the same number of total importations as the posterior median. Within this analysis, only sequences with unambiguous postcode districts were used, resulting in a dataset of 25,139 sequences for the seven largest transmission lineages and 24,411 across 280 smaller lineages, which were extracted from the master COG-UK alignment, described in “Genomic Data” above. Within those postcode districts, we assigned random coordinates to each sequence, as the continuous phylogeographic analysis does not permit identical values. This was achieved using geographical data from ^87^. We then reconstructed the geographic movement of nodes on a fixed tree (pruned from the overall MCC tree) in BEAST v.1.10^81^, using a relaxed random walk (RRW) model^88^, and a Cauchy distribution to account for among-branch heterogeneity in dispersal velocity. Large lineages were inferred independently, and all small lineages were inferred in a single run, with the shared parameters for likelihood, precision, and covariance of coordinates, but independent estimates of diffusion rate and trait likelihood. Following this run, 22 small introductions were removed due to their chains not converging to the same posterior. An MCC tree was then generated using TreeAnnotator^81^ to summarise the posterior tree distribution for all lineages. Visualisations were made using a custom Python script. XML files were generated using beastgen.py (https://github.com/ViralVerity/beastgenpy), and can be found along with data processing and visualisation scripts on GitHub.

For the export analyses we compare Greater London to Greater Manchester which consists of the UTLAs Salford, Trafford, Stockport, Oldham, Bolton, Tameside, Bury, Rochdale, Wigan and Manchester.

#### State level incidence data from India

State level COVID-19 case count data were extracted from https://api.covid19india.org/csv/latest/states.csv.

#### Incidence data from England

COVID-19 case count data for each Local Tier Local Authority were downloaded via https://coronavirus.data.gov.uk/details/download.

### Travel history data

Four sources of data were compiled to provide the travel history for laboratory confirmed cases, depending on availability for each individual case: (1) public health passenger locator forms are required for entry into the UK; (2) routine public health contact tracing data including UK Health Security Agency Second Generation Surveillance System (SGSS)^89^, (3) COVID-19 test requests with reported travel associations and (4) responses to additional telephone interviews for cases.

### Covariate processing for statistical analyses

Country-level COVID-19 case count and vaccination data from 1st January 2020 to 9th July 2021 were downloaded via Our World in Data https://github.com/owid/covid-19-data/blob/master/public/data/owid-covid-data.csv. The number of individuals who have received a partial course of the vaccine per day by country was obtained from the difference in partially vaccinated individuals from consecutive days. The same operation was used to obtain the number of new fully vaccinated individuals per day by country . To deal with missing values, we assumed the vaccination rate to be constant between any two closest dates with vaccination data. This assumption was only applied when the time period between successive vaccination data entries was less than 7 days. Missing vaccination data for more than 6 consecutive days resulted in all of the new vaccinations administered from the last entry date to the next entry date to have been administered on the next entry date.

#### COVID-19 case count and vaccination data for the United Kingdom

COVID-19 case count data and vaccination data were downloaded by UTLA from 30th January 2020 to 28th July 2021 by specimen and dosage date respectively via https://coronavirus.data.gov.uk/details/download. These data include positive lab-based polymerase chain reaction (PCR) tests and positive LFT tests, but do not include tests where the LFT was positive and PCR follow up tests were negative (see more details here^90^). COVID-19 case count at the United Kingdom country level was calculated by aggregating case data on the UTLA-level. Additionally, to match the genomic data, the COVID-19 case count and vaccination data for some UTLAs were aggregated under an area code made up of these multiple UTLAs (see Table S3). All entries with the recently discontinued area code ‘E10000002’ were assigned the new area code ‘E06000060’.

#### United Kingdom population data

UTLA-level 2020-mid-year population size estimates were downloaded via https://www.ons.gov.uk/peoplepopulationandcommunity/populationandmigration/populationestimates/datasets/populationestimatesforukenglandandwalesscotlandandnorthernireland#:~:text=Mid-2020%20edition%20of%20this%20dataset%202021%20local%20authority%20boundaries. Population size data were used to calculate the proportion of the population that was partially or fully vaccinated in a location.

#### State level COVID-19 case count data from the U.S

For U.S. states, COVID-19 case count data from 22nd January 2020 to 12th July 2021 were downloaded via https://data.cdc.gov/Case-Surveillance/United-States-COVID-19-Cases-and-Deaths-by-State-o/9mfq-cb36. Vaccination data from 14th December 2020 to 12th July 2021 were downloaded via https://data.cdc.gov/Vaccinations/COVID-19-Vaccinations-in-the-United-States-Jurisdi/unsk-b7f. The number of new partially vaccinated individuals per day by state was calculated from the difference in total partially vaccinated individuals from consecutive days. The same operation was used to obtain the number of new fully vaccinated individuals per day by state .

#### U.S. states population level immunity

Daily population immunity estimates for COVID-19 was downloaded by the U.S. state from 26th January 2021 to 9th June 2021 via https://popimmunity.biosci.gatech.edu/. A sampling bias of four was selected for (i.e. a sampling fraction of 25% is assumed) using fully vaccinated individuals for the calculation of the estimate^91^. For our analysis at the weekly level, the mean of the week’s daily estimated population immunity was calculated for each state.

#### State level population data from U.S

The most recent population size estimate for each US state for the year 2019 was downloaded via https://www.census.gov/data/datasets/time-series/demo/popest/2010s-state-total.html.

#### Global population data

Country-level population size estimates for the year 2021 were downloaded via https://data.worldbank.org/indicator/SP.POP.TOTL?name_desc=false.

#### Aggregated and anonymised human mobility data

We used the Google COVID-19 Aggregated Mobility Research Dataset described in detail in^47,92^, which contains anonymized relative mobility flows aggregated over users who have turned on the *Location History* setting, which is turned off by default. This is similar to the data used to show how busy certain types of places are in Google Maps — helping identify when a local business tends to be the most crowded. The mobility flux is aggregated per week, between pairs of approximately 5km^2^ cells worldwide, and for the purpose of this study further aggregated for LTLAs in the United Kingdom (https://geoportal.statistics.gov.uk/datasets/lower-tier-local-authority-to-upper-tier-local-authority-december-2016-lookup-in-england-and-wales/explore), U.S. states (https://gadm.org/), and country level (https://gadm.org/) for all other countries for the time period of October 29th, 2020 to June 6th, 2021.

To produce this dataset, machine learning is applied to log data to automatically segment it into semantic trips. To provide strong privacy guarantees^93^, all trips were anonymized and aggregated using a differentially private mechanism to aggregate flows over time (see https://policies.google.com/technologies/anonymization). This research is done on the resulting heavily aggregated and differentially private data. No individual user data was ever manually inspected, only heavily aggregated flows of large populations were handled. All anonymized trips are processed in aggregate to extract their origin and destination location and time. For example, if users travelled from location a to location b within time interval t, the corresponding cell (a,b,t) in the tensor would be n∓err, where err is Laplacian noise. The automated Laplace mechanism adds random noise drawn from a zero mean Laplacian distribution and yields (∊, δ)-differential privacy guarantee of ∊ = 0.66 and δ = 2.1 × 10−29 per metric. Specifically, for each week W and each location pair (A,B), we compute the number of unique users who took a trip from location A to location B during week W. To each of these metrics, we add Laplace noise from a zero-mean distribution of scale 1/0.66. We then remove all metrics for which the noisy number of users is lower than 100, following the process described in ^93^, and publish the rest. This yields that each metric we publish satisfies (ε,δ)-differential privacy with values defined above. The parameter ∊ controls the noise intensity in terms of its variance, while δ represents the deviation from pure ∊-privacy. The closer they are to zero, the stronger the privacy guarantees.

These results should be interpreted in light of several important limitations. First, the Google mobility data is limited to smartphone users who have opted in to Google’s *Location History* feature, which is off by default. These data may not be representative of the population as whole, and furthermore their representativeness may vary by location. Importantly, these limited data are only viewed through the lens of differential privacy algorithms, specifically designed to protect user anonymity and obscure fine detail. Moreover, comparisons across rather than within locations are only descriptive since these regions can differ in substantial ways.

#### Flight data

We used data from the International Air Transport Association (IATA)^94^ on the monthly number of confirmed passengers on flights (direct and indirect) from India to all other countries from January 2021 to June 2021.

#### Estimated Importation Intensity (EII)

We estimated the weekly importation intensity of the Delta variant for each destination location at the weekly level using the human mobility, GISAID and COG-UK genomic data and COVID-19 case data. An importation intensity value was calculated for each international movement by multiplying the proportion of Delta in the location of origin, the total number of new weekly reported COVID-19 cases and the movement intensity between each origin location and the destination location. We then aggregated all importation intensity values by week and destination location to obtain the EII.

#### Estimated Exportation Intensity (EEI)

We estimated the exportation intensity of the Delta variant for each location of origin at the weekly level using aggregated human mobility, genomic and case count data. An exportation intensity value was calculated for each international movement by multiplying the proportion of Delta in the country of origin, the total number of new weekly reported cases and the movement intensity between the country of origin and the destination country. We then aggregated all importation intensity values by week and origin location to obtain the EEI.

#### Estimated local human mobility intensity

To obtain an estimate of the intensity of human mobility within a location, we calculated a ’relative self-mobility’ value indicating the intensity of mobility within a location (where the origin and destination of the trips are the same) as a percent of the highest recorded of movement within this location in our mobility data during the time period from 2020-03-22 to 2021-06-06 using the human mobility data described above.

#### New Delta lineage introductions

Daily new lineage introductions into the United Kingdom by UTLA were obtained from the continuous phylogenetic analysis described above. The data were aggregated by week and UTLA.

### Statistical modelling of Delta growth

Data pre-processing: we kept data starting from the 13th (week commencing 28th March 2021) and 11th (week commencing 14th March 2021) epidemiological weeks for England and the USA, respectively. These dates are referred to as baseline elsewhere in the main text. We excluded weeks after the first time 95% of samples were observed to be Delta in each UTLA (England) or state (USA) because after this point we can no longer estimate the relative growth rates reliably since Delta saturation has been reached. Therefore, each UTLA or state potentially had a different number of time points (Extended Data Table 8). Finally, we kept only those UTLAs or states which have data on Delta for at least three weeks (which are not required to be consecutive). In the final dataset for England and the USA, we had 590 (66 UTLAs and 8 weeks on average) and 735 (51 states and 14 weeks on average) and observations, respectively (Extended Data Fig. 8).

#### Model

In what follows, we model the dynamics of Delta penetration within a UTLA or state: we refer to these levels of spatial unit as *subregions*. Here, we model how the number of Delta samples per subregion, i, varies over time, t (here, measured in weeks). The background transmission conditions driving the observed number of delta samples in a given subregion may be similar to the subregions within the same region. As such, we model this variation hierarchically and index variables at the subregional level by *i*[*j*] to indicate that subregion *i* is nested within region *j*: in England, regions correspond to NUTS1 units and, in the USA, to units also named regions. We use a binomial sampling distribution to model the number of Delta samples 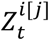,

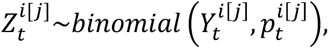

where 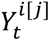 is the total number of sequenced samples, and 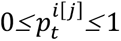 is the corresponding proportion of Delta samples in subregion *i* in week *t*. We then transform this probability, so that it is on the (unconstrained) logit scale:

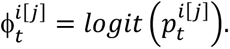

A key quantity of interest is the *relative growth* in the proportion of Delta on the logit (i.e. log-odds) scale, which we estimate weekly and is denoted by 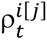, where

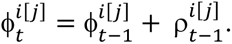

Relative growth for each subregion, *i*, is modelled spatially as depending hierarchically on its containing region, *j*. It is also assumed to depend on subregion-specific covariates:

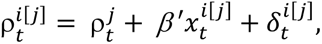

where 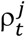 is a region-level growth trend, 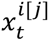 is a vector of covariates, and 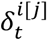 is a subregion- and week-specific term representing the deviation from the region-level growth. To account for temporal autocorrelation in the relative growth rate, a given region’s relative growth is assumed to follow a random walk centred around its relative growth in the previous week:

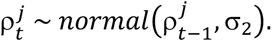

We chose to use different sets of covariates in our chosen “best” models for England and the USA. These covariates were chosen as important predictors if including them in the model improved the model fit (as indicated by higher log likelihood; Extended Data Table 4), gave better out of sample prediction (Extended Data Table 6), and if they were confounding variables. For England, the covariates included relative self-mobility and time since baseline (in weeks, standardized by subtracting the mean and dividing by the standard deviation); for the USA, it included baseline immunity (natural infection or vaccination induced immunity), relative self-mobility, and time since baseline. Including data on importations decreased the number of observations due to missing data on importations from 735 to 387 in the USA (using Estimated Importation Intensity, which was standardized before including in the regression) and from 590 to 299 in England (using New Delta lineage introductions, which was square root transformed because of skewed positive data). The effect size (95% credible interval) of importations was negligible when added to the “best” model: 0.00(−0.14, 0.14) for USA and -0.04(−0.07, -0.01) for England, and hence importation was not included in the final models.

We estimated our model in a Bayesian framework and chose priors (Extended Data Table 9) so that a wide range of possible Delta proportions were possible yet were centred on low values in the absence of further information: our prior predictive distributions in Figure S11b and Figure S14b illustrate these characteristics.

The computations were done using R and Stan using four parallel chains with 20,000 to 40,000 iterations (depending on the model), half of which were discarded as warmup iterations; the chains were subsequently thinned by a factor of 10. In all cases, MCMC sampling was diagnosed as converged with 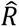 < 1.01, and bulk and tail effective sample sizes >400 for all parameters. For the England model with no variables when used for model comparison, we obtained Rhat < 1.01 and bulk ESS > 400 for all parameters but there were 284 out of 4,410 parameters where tail ESS < 400 even with 40,000 iterations (minimum tail ESS = 169.6). In this model the last two weeks were held-out from each UTLA to perform out of sample predictions, resulting in a smaller dataset. This could be the reason for difficulty in convergence with 40,000 iterations.

Our model outputted two sets of key quantities: the weekly relative growth rate of Delta over time 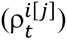 and the estimated “effect” of a variable on Delta growth (*β*). To determine the implications of the effect sizes, we computed the estimated proportion of Delta samples when the covariates took factual versus counterfactual values. We considered counterfactual scenarios for relative self-mobility in England and baseline immunity in the USA, holding all other covariates at their factual values. The counterfactual scenarios we considered were:

`o` England: “Minimum mobility” (relative self-mobility = 0), “Maximum mobility” (relative self-mobility = 1)
`o` USA: No prior immunity (baseline immunity = 0), 90% people immune at baseline (baseline immunity = 0.90)

Simulation and model robustness: To test model parameter identifiability, we performed inference on simulated data. We fixed the parameters and simulated from the model to create hypothetical data (with 5 regions, each with 6 sub-regions (i.e. UTLA or state) and 15 time points). We then used these data to estimate the known parameters. We were reasonably able to recover our parameters and the model converged with R <1.01, bulk and tail effective sample sizes >400 after 20,000 iterations, discarding 10,00 warm-up iterations and thinning by a factor of 10 (Table S7 and Figure S15).

## Acknowledgements

COG-UK is supported by funding from the Medical Research Council (MRC) part of UK Research & Innovation (UKRI), the National Institute of Health Research (NIHR) [grant code: MC_PC_19027], and Genome Research Limited, operating as the Wellcome Sanger Institute. M.U.G.K. acknowledges support from a Branco Weiss Fellowship, Google.org, and The Rockefeller Foundation. S.D. and M.U.G.K. acknowledge support from the European Union Horizon 2020 project MOOD [grant agreement number 874850]. O.G.P., M.U.G.K., L.dP., and A.E.Z. acknowledge support from the Oxford Martin School. V.H. was supported by the Biotechnology and Biological Sciences Research Council (BBSRC) [grant number BB/M010996/1]. S.D. is supported by the Fonds National de la Recherche Scientifique (FNRS, Belgium). J.T.M, R.C. and A.R. acknowledge support from the Wellcome Trust [Collaborators Award 206298/Z/17/Z - ARTIC network]. A.R. is also supported by the European Research Council [grant agreement number 725422 - ReservoirDOCS] and Bill & Melinda Gates Foundation [OPP1175094 – HIV-PANGEA II]. C.R. was supported by a Fondation Botnar Research Award (programme grant 6063). G.B. acknowledges support from the Research Foundation - Flanders (Fonds voor Wetenschappelijk Onderzoek-Vlaanderen, GOE1420N and G098321N) and from the Interne Fondsen KU Leuven/Internal Funds KU Leuven under grant agreement C14/18/094. A.OT is supported by the Wellcome Trust Hosts, Pathogens & Global Health Programme [grant number: grant.203783/Z16/Z] and Fast Grants [award number: 2236]. SB is supported by the Clarendon Scholarship, University of Oxford and NERC DTP [grant number NE/S007474/1]. M.A.S. acknowledges support from US National Institutes of Health grant R01 AI153044. X.J. acknowledges support from US National Institutes of Health grant U19 AI135995. T.P.P and W.S.B. acknowledge support from the G2P-UK National Virology Consortium funded by the MRC [MR/W005611/1]. IIB is supported by the Canadian Institutes of Health Research [grant 02179-000]. The contents of this publication are the sole responsibility of the authors and do not necessarily reflect the views of the European Commission or any of the other funders.

## Author contributions

J.T.M., V.H., S.B., O.G.P., A.R., M.U.G.K. conceived and planned the research. J.T.M., V.H., S.B., R.E.P., R.I., C.R., C.H., I.I.B., O.G.P., A.R., M.U.G.K. analysed and processed the data. S.B. performed statistical epidemiological analyses. B.C.L., M.U.G.K., E.V., S.Bhatt, S.D., G.B., X.J. and M.A.S. advised on statistical methodology. J.T.M., V.H., B.J., R.C., A.O’T., N.J.M., D.A., A.R., designed and implemented genomic data processing pipelines. J.T.M., V.H., O.G.P., S.B., R.E.P., A.R., M.U.G.K. wrote the first draft of the manuscript. All authors contributed to editing and interpreting the results. A.R., O.G.P., and M.U.G.K. jointly supervised the work.

## Correspondence and requests for materials

should be addressed to O.G.P, A.R. or M.U.G.K.

## Extended Data Tables

**Extended Data Table 1:** Delta variant mutations compared to reference Wuhan-1.

| Mutation |
| --- |
| ORF1a:A1306S |
| ORF1a:P2046L |
| ORF1a:P2287S |
| ORF1a:V2930L |
| ORF1a:T3255I |
| ORF1a:T3646A |
| ORF1b:P314L |
| ORF1b:G662S |
| ORF1b:P1000L |
| ORF1b:A1918V |
| S:T19R |
| S:G142D |
| S:E156G |
| S:157/158del |
| S:L452R |
| S:T478K |
| S:D614G |
| S:P681R |
| S:D950N |
| ORF3a:S26L |
| M:I82T |
| ORF7a:V82A |
| ORF7a:T120I |
| ORF7b:T40I |
| ORF8:S84L |
| ORF8:119/120del |
| N:D63G |
| N:R203M |
| N:G215C |
| N:D377Y |

**Extended Data Table 2:** Table showing the percentage of cases sequenced in each state in India during the study period between the 28th of November 2020 to the 16th of May 2021 (also see Fig. S11).

| State | Number of Cases | Number of Genomic Sequences | Fraction of Cases Sequenced (%) |
| --- | --- | --- | --- |
| Andhra Pradesh | 568,428 | 468 | 0.08 |
| Bihar | 417,356 | 42 | 0.010 |
| Chandigarh | 38,121 | 16 | 0.042 |
| Chhattisgarh | 677,752 | 144 | 0.021 |
| Delhi | 832,125 | 902 | 0.11 |
| Goa | 88,167 | 34 | 0.039 |
| Gujarat | 545,905 | 841 | 0.15 |
| Haryana | 463,714 | 534 | 0.12 |
| Himachal Pradesh | 121,263 | 24 | 0.020 |
| Jammu and Kashmir | 135,225 | 51 | 0.038 |
| Jharkhand | 206,716 | 242 | 0.12 |
| Karnataka | 1,320,854 | 183 | 0.014 |
| Ladakh | 8,124 | 22 | 0.27 |
| Madhya Pradesh | 528,154 | 88 | 0.017 |
| Maharashtra | 3,563,937 | 1,169 | 0.033 |
| Odisha | 294,435 | 414 | 0.14 |
| Puducherry | 47,604 | 128 | 0.27 |
| Punjab | 346,900 | 122 | 0.035 |
| Sikkim | 6,443 | 28 | 0.43 |
| Tamil Nadu | 819,170 | 961 | 0.12 |
| Telangana | 260,405 | 724 | 0.28 |
| Tripura | 8,175 | 116 | 1.14 |
| Uttar Pradesh | 1,079,746 | 485 | 0.045 |
| Uttarakhand | 213,335 | 210 | 0.098 |
| West Bengal | 655,984 | 906 | 0.14 |

**Extended Data Table 3:** Grouping of Upper Tier Local Authority area codes under higher level area codes as used in the analyses.

| Area and Area Code | Constituents |
| --- | --- |
| Greater London,<br>E13000001\|E13000002 | E09000007, E09000011, E09000012, E09000013, E09000019, E09000020, E09000022, E09000023, E09000028, E09000030, E09000032, E09000033, E09000001, E09000002, E09000003, E09000004, E09000005, E09000006, E09000008, E09000009, E09000010, E09000014, E09000015, E09000016, E09000017, E09000018, E09000021, E09000027, E09000024, E09000025, E09000026, E09000029, E09000031 |
| West Midlands,<br>E11000005 | E08000026, E08000029, E08000025, E08000028, E08000030, E08000027, E08000031 |
| South Yorkshire,<br>E11000003 | E08000019, E08000018, E08000017, E08000016 |
| Tyne and Wear,<br>E11000007 | E08000037, E08000021, E08000022, E08000023, E08000024 |
| Merseyside, E11000002 | E08000012, E08000014, E08000011, E08000013, E08000015 |
| Greater Manchester,<br>E11000001 | E08000003, E08000007, E08000008, E08000004, E08000005, E08000002, E08000001, E08000010, E08000006, E08000009 |
| West Yorkshire,<br>E11000006 | E08000035, E08000036, E08000034, E08000033, E08000032 |

**Extended Data Table 4:** Parameter estimates of covariates in the model estimating relative growth over time.

| Country | Covariate | Posterior mean (95% Bayesian credible interval) |
| --- | --- | --- |
| US States | Baseline immunity | 0.60 (0.12, 1.13) |
|  | Relative self mobility | -0.08 (-0.68, 0.47) |
|  | Time since baseline (weeks) | 0.02 (-0.04, 0.10) |
| England | Relative self mobility | 0.43 (-0.08, 1.00) |
|  | Time since baseline (weeks) | 0.01 (-0.03, 0.04) |

**Extended Data Table 5:** Parameter estimates of covariates in the model (with natural infection induced immunity only) estimating relative growth over time.

| Country | Covariate | Posterior mean (95% Bayesian credible interval) |
| --- | --- | --- |
| US States | Baseline immunity (natural infection induced) | 0.59 (0.09, 1.07) |
|  | Relative self mobility | -0.04 (-0.57, 0.49) |
|  | Time since baseline (weeks) | 0.02 (-0.04, 0.09) |

**Extended Data Table 6:**
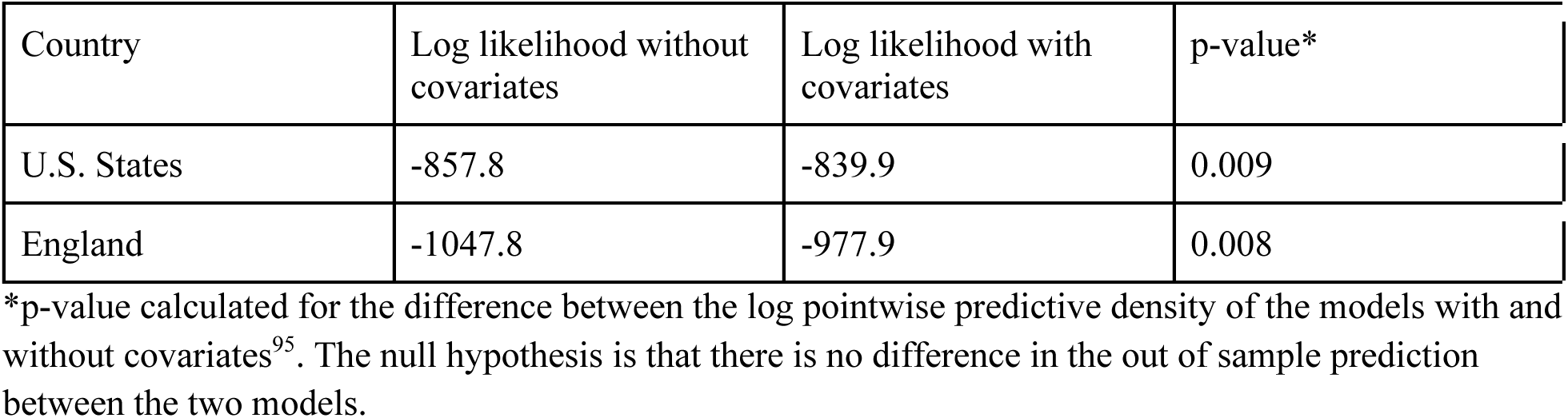
Out of sample prediction (leaving out 20% of the data i.e. last three and last two weeks for USA and England, respectively) comparing models with and without covariates (see details on covariates in Extended Data Table 4).

**Extended Data Table 7:** Simulation: Known vs estimated parameters.

| Parameter (example covariate) | Known value | Posterior mean (95% Bayesian credible interval) |
| --- | --- | --- |
| Beta1 (baseline immunity) | 2.0 | 1.67 (-0.43, 3.62) |
| Beta2 (time) | 0.1 | 0.18 (0.13, 0.23) |
| sigma1 | 0.6 | 0.49 (0.43, 0.55) |
| sigma2 | 0.2 | 0.17 (0.09, 0.27) |

**Extended Data Table 8:** Table describing the data at the sub-regional level for England and USA as mean (minimum, maximum) after data pre-processing.

|  | England | USA |
| --- | --- | --- |
| # data points (weekly) | 8.9 (3, 11) | 14.4 (12, 15) |
| % Delta samples per data point | 22.7% (0%, 94.9%) | 7.2% (0%, 91.2%) |
| # samples per data point | 118.4 (1, 2714) | 396.1 (1, 3550) |
| Relative self-mobility | 0.8 (0.4, 1) | 0.9 (0.3, 1) |
| Baseline immunity (proportion of individuals with natural infection or vaccination induced immunity) | NA | 0.4 (0.2, 0.6) |

**Extended Data Table 9:** Prior distributions for model parameters.

| Parameter | Prior distribution |
| --- | --- |
| $\rho_1^j$ | normal (0,1) |
| $\delta_t^{i[j]}$ | normal (0, $\sigma_1$) |
| $\phi_1^{i[j]}$ | normal ($a^j, b^j$) |
| $a^j$ | half-normal (−3,1) |
| $b^j$ | half-normal (1.5,1) |
| $\sigma_1$ | half-normal (0,5) |
| $\sigma_2$ | half-normal (0,5) |
| $\beta$ | half-normal (0,5) |

## Extended Data Figures

**Extended Data Figure 1:**
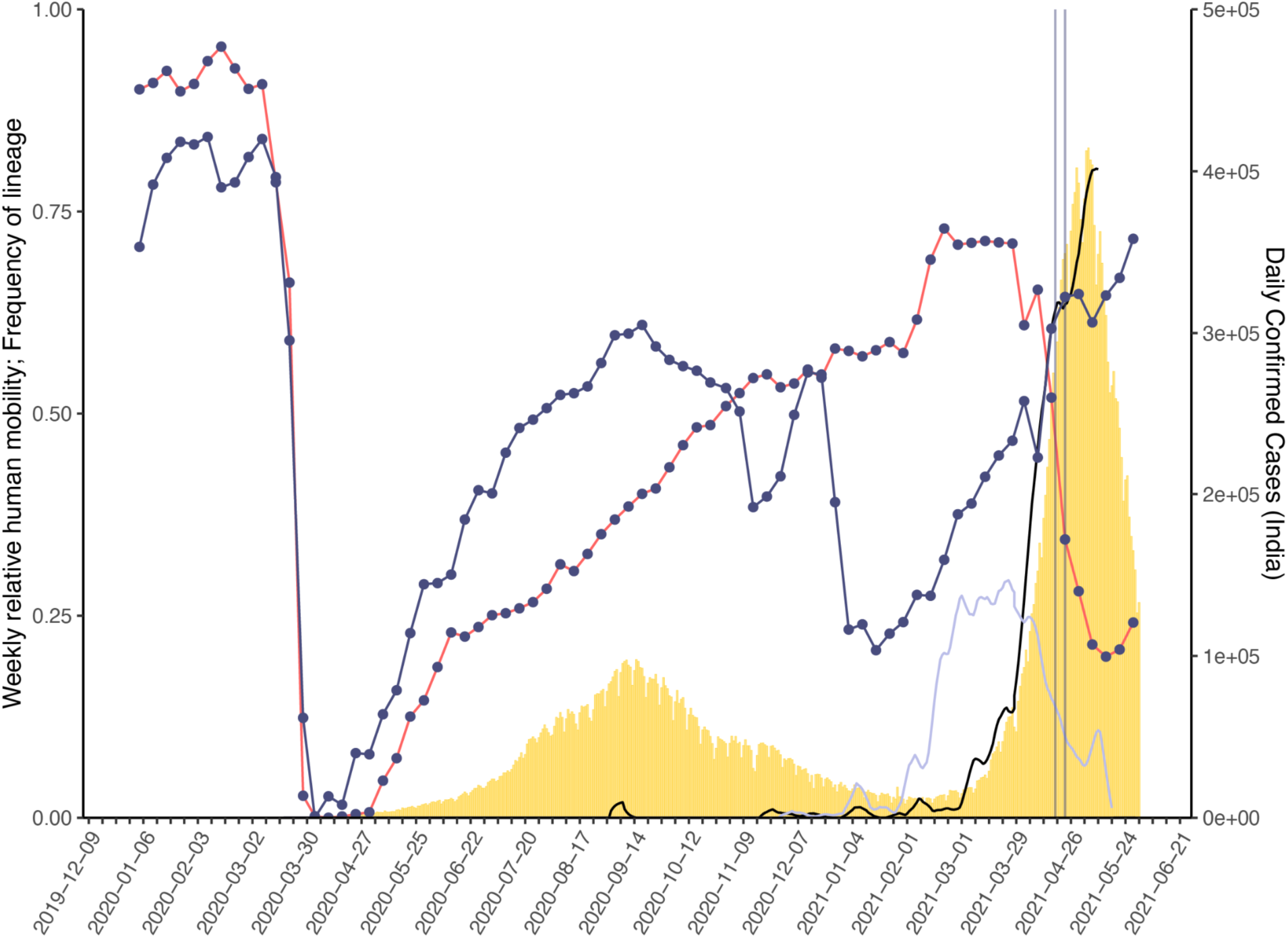
Daily number of reported SARS-CoV-2 cases (yellow bars, right hand axis) in India. Weekly human movements in England, relative to the maximum in England (dark blue line, left hand axis, Methods), and in India, relative to the maximum in India (red line, left hand axis, Methods). Proportion of genomes in India that are assigned to lineages B.1.617.2 (black line, no points) and B.1.617.1 (light blue line, no points) (left hand axis). First vertical line represents the announcement of the quarantine policy for arrivals of travellers from India to England (17 March 2021) and the second vertical line represents the date of implementation (23 March 2021).

**Extended Data Figure 2:**
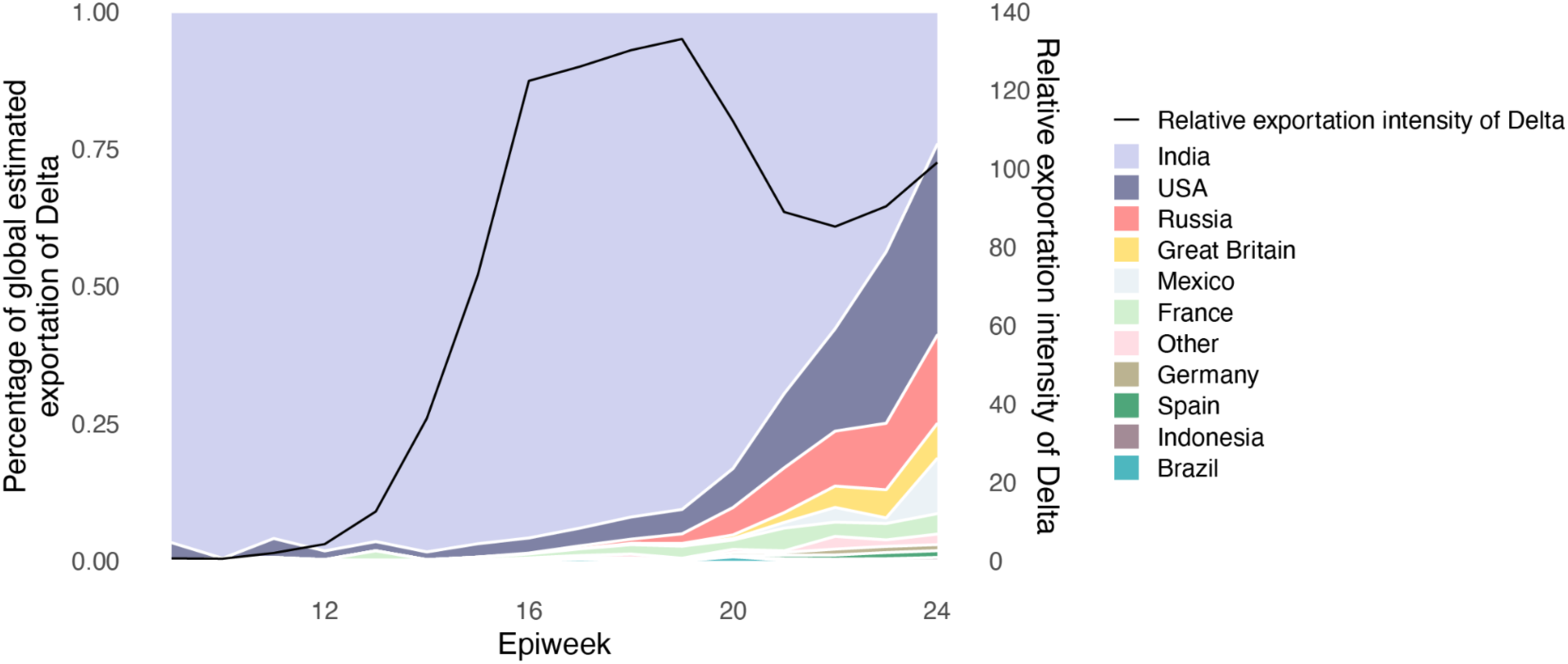
Proportion of weekly Estimated Exportation Intensity (EEI) of Delta by country. See Methods for details of calculation (left y-axis). The black line represents the total EEI by week (right y-axis).

**Extended Data Figure 3:**
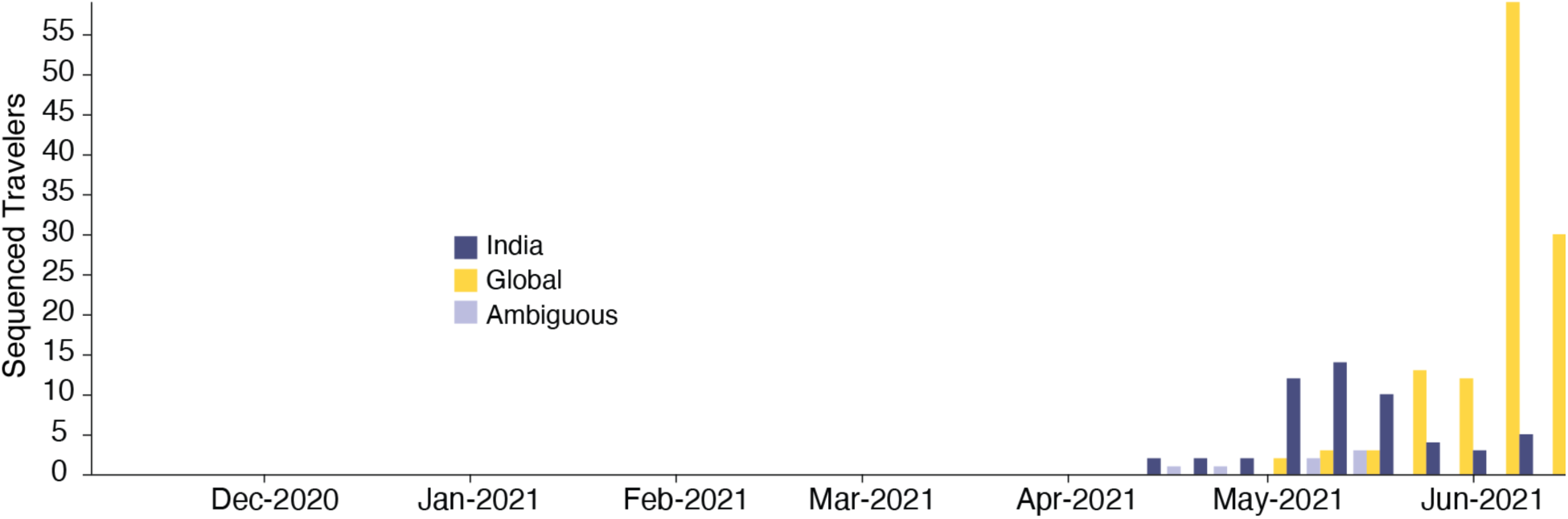
Temporal distribution of genomic isolates from the AY.4 sublineage with travel history, by the likely location of exposure.

**Extended Data Figure 4:**
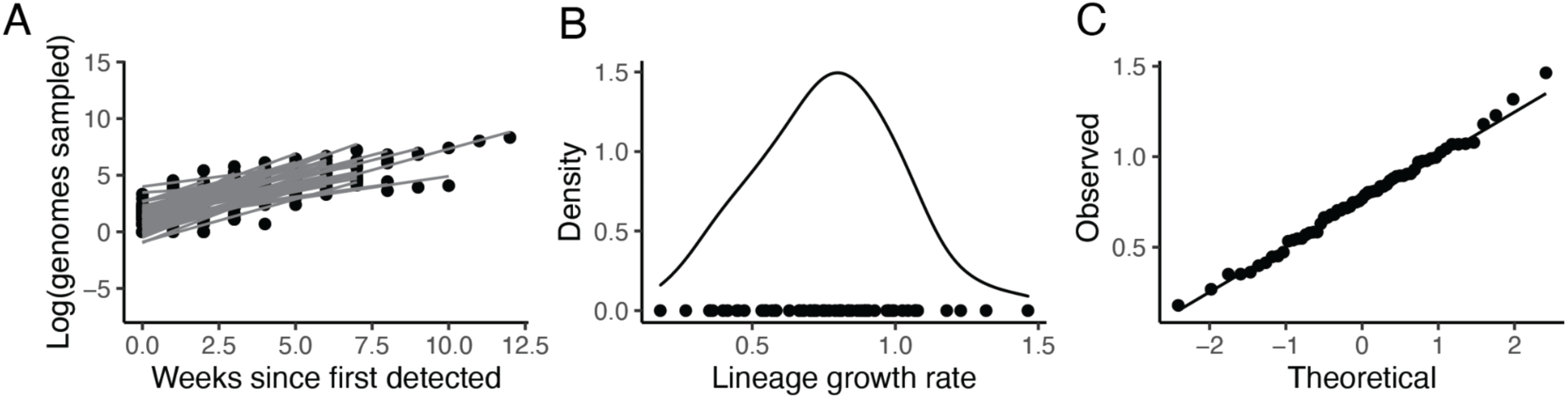
Growth of transmission lineages in England for lineages observed for at least 3 weeks and with >100 genomes sampled in total. A) The log number of weekly sampled genomes per transmission lineage plotted over time. Lines represent a linear fit (assuming exponential growth). B) Distribution of growth rates (slopes in A). C) Quantile Quantile plot comparing the observed quantiles in the growth rate distribution to theoretical quantiles from a normal distribution.

**Extended Data Figure 5:**
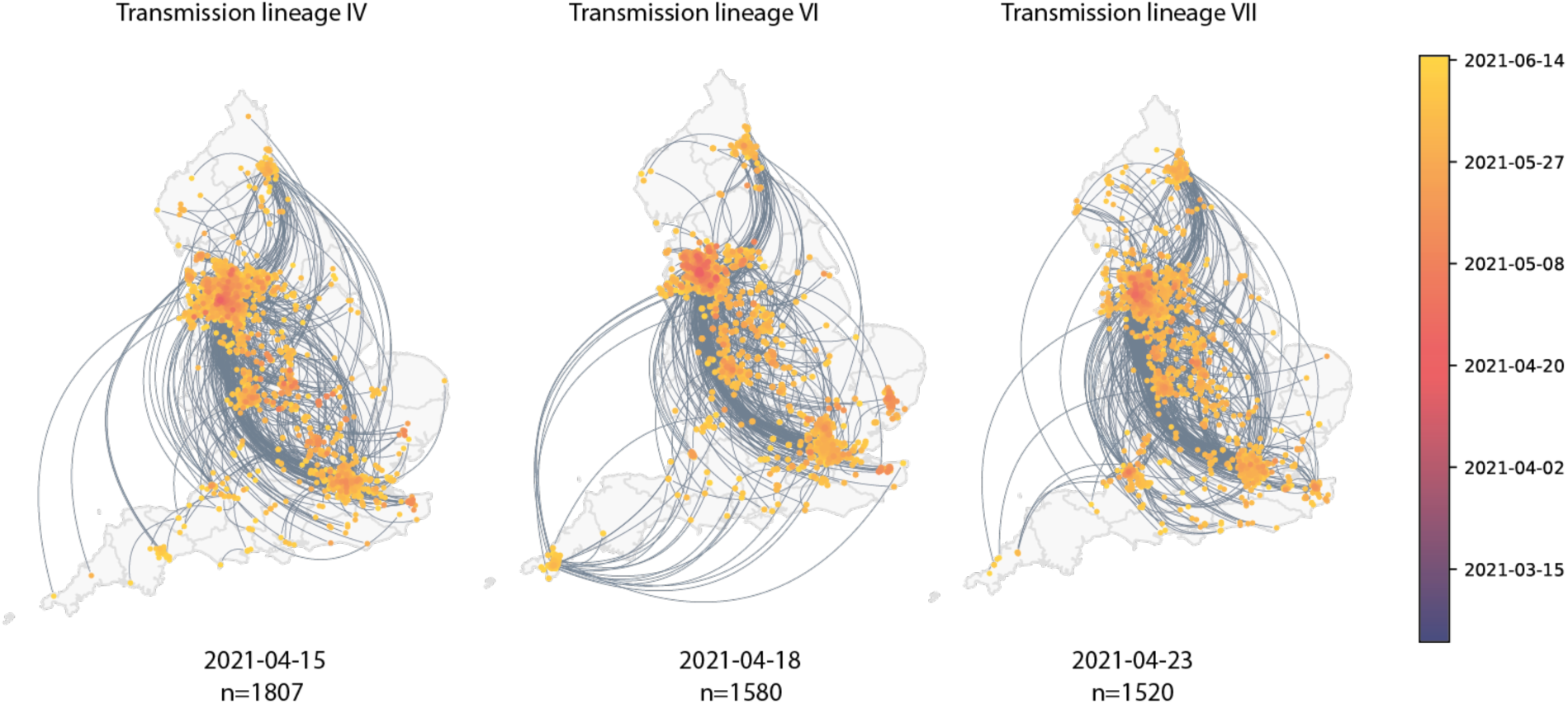
Maps showing virus movements inferred using continuous phylogeographic analysis for the fourth, sixth and seventh largest transmission lineages. Direction of movement is anti-clockwise, and dots are coloured by date.

**Extended Data Figure 6:**
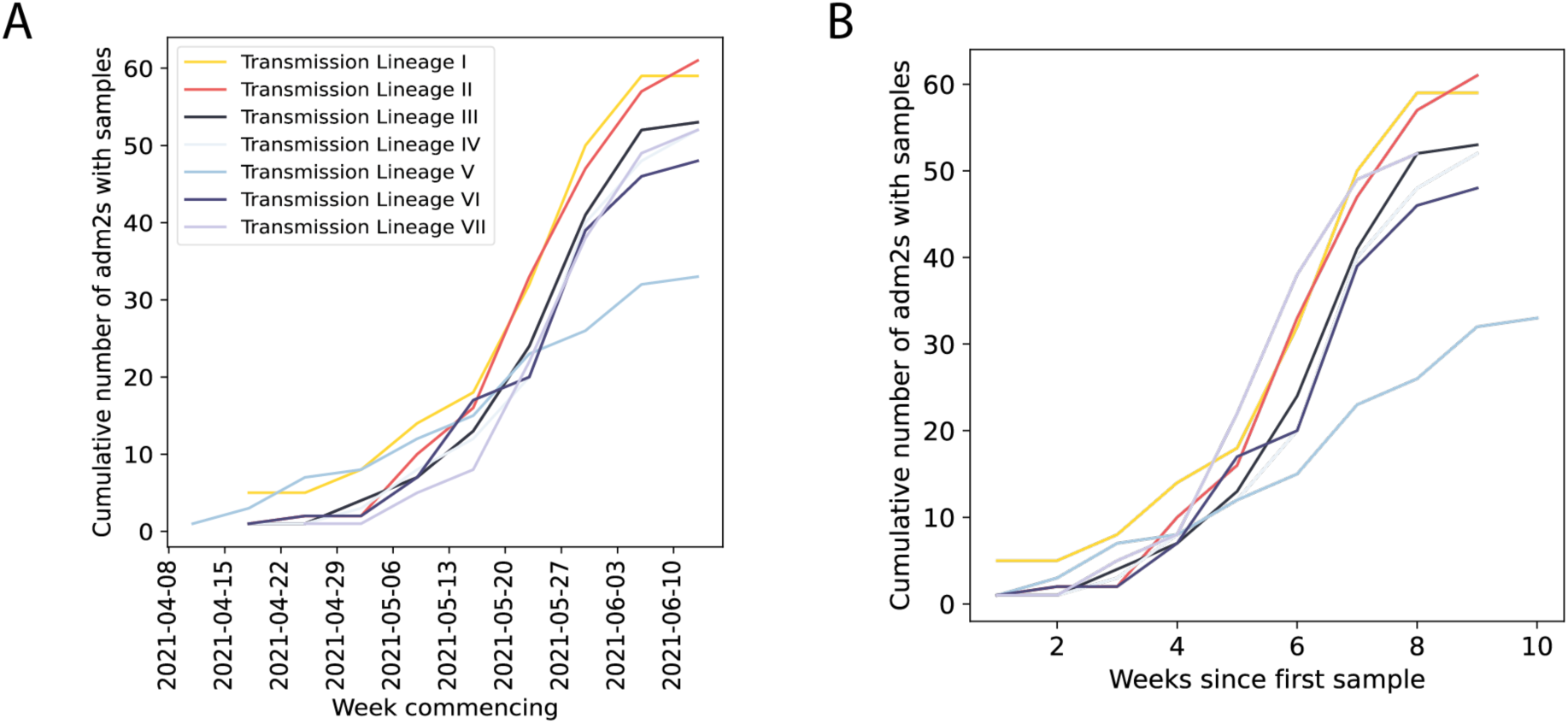
Cumulative number of UTLAs that the five largest Delta transmission lineages are sampled in absolute (A) and relative (B) time.

**Extended Data Figure 7:**
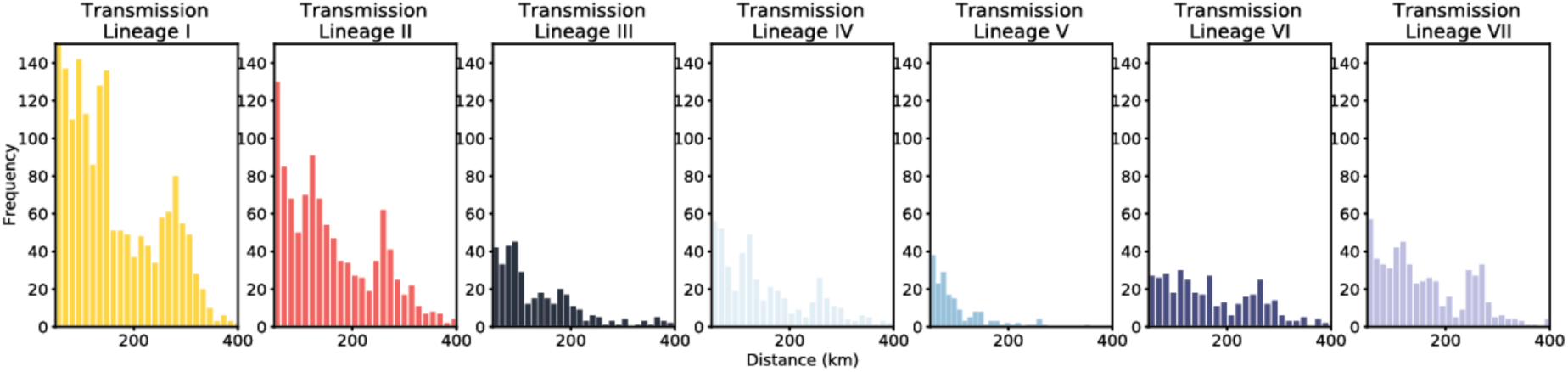
Histograms of the distance of viral movements over 50km for each of the largest seven Delta transmission lineages in England.

**Extended Data Figure 8:**
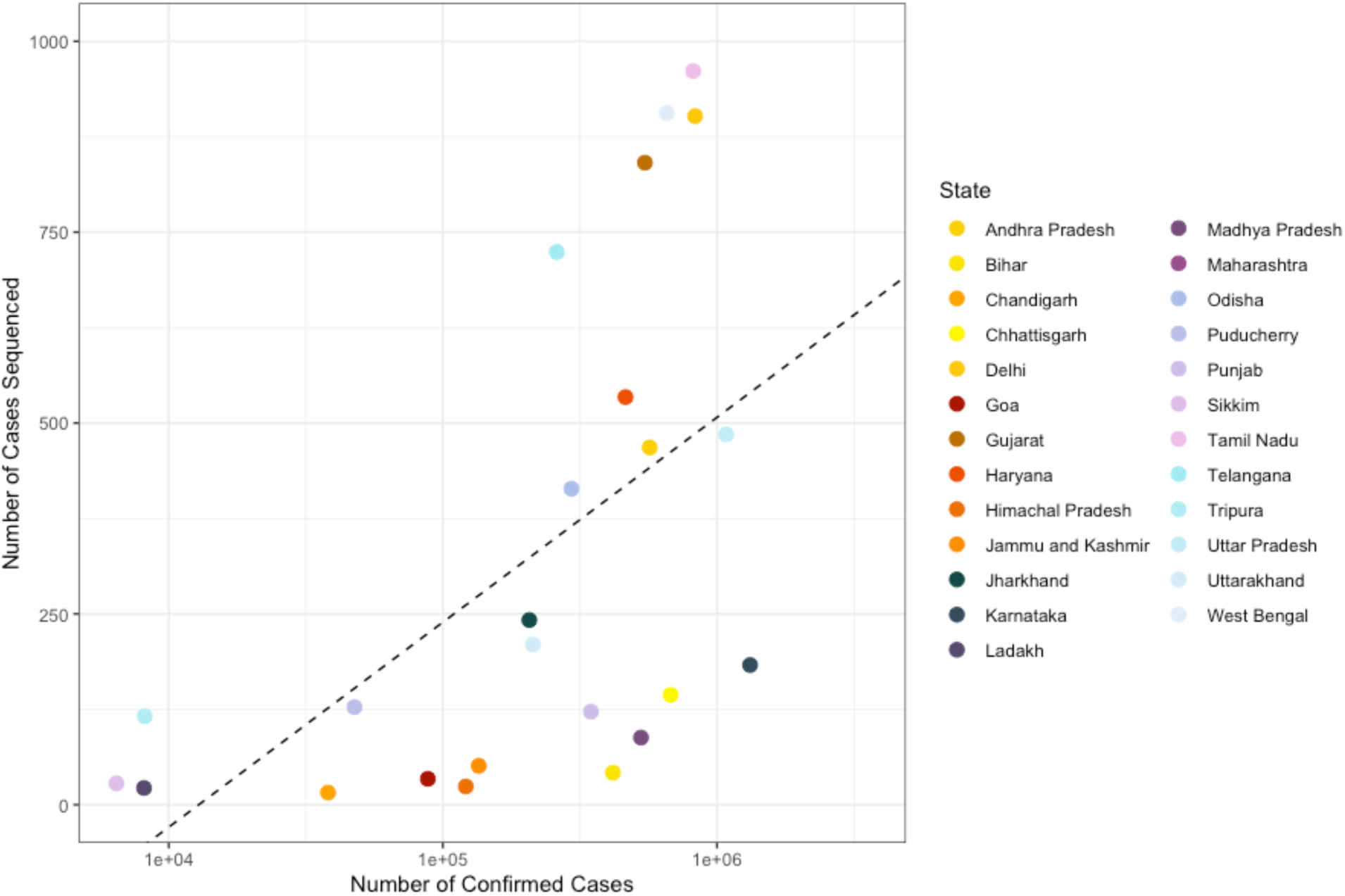
Scatter plot showing the number of confirmed cases per state in India vs. the number of cases sequenced in that state in India between 28th of November 2020 to the 16th of May 2021. In states above the line more than the mean number of cases were sequenced.

**Extended Data Figure 9:**
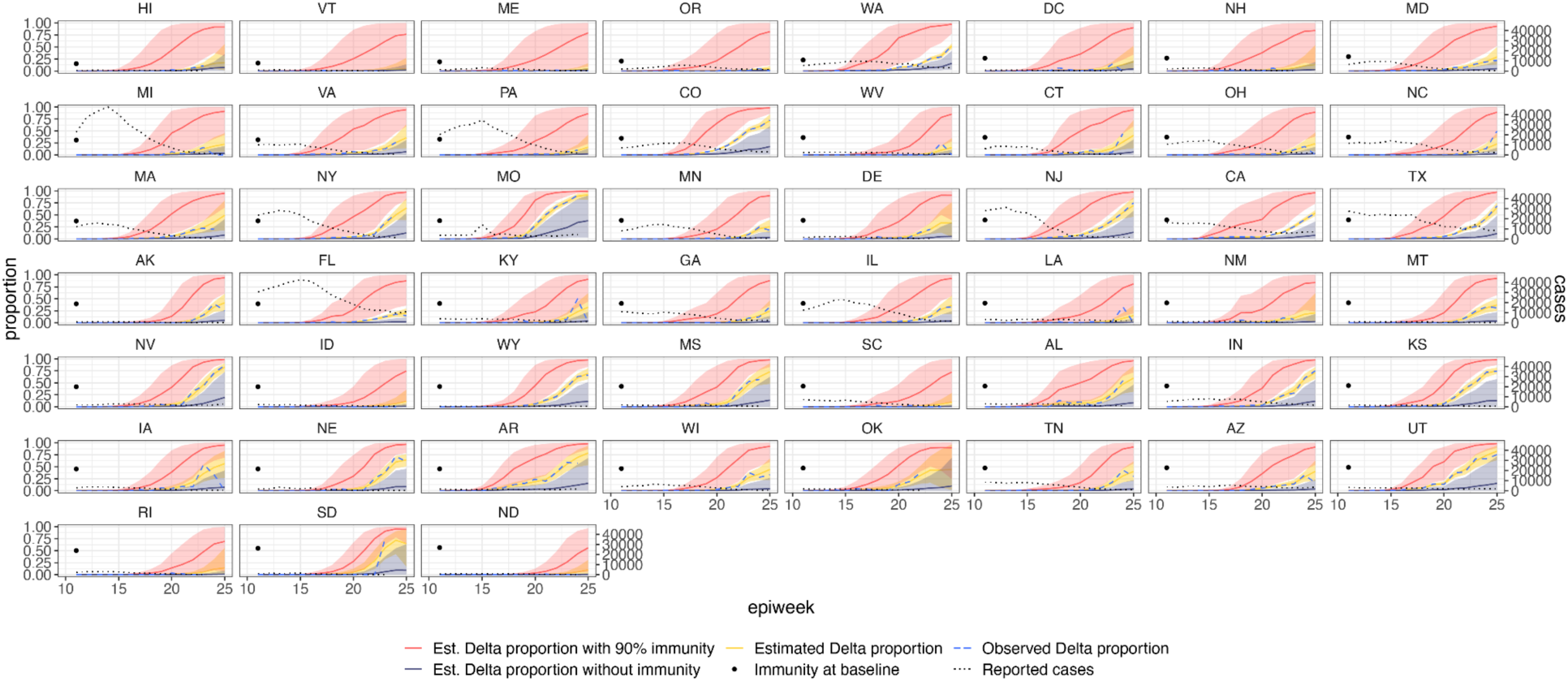
Estimated and observed proportions of Delta variant samples across US states (yellow and blue dashed respectively), counterfactual scenarios: no prior immunity (purple), 90% immunity from the beginning (red), and reported number of cases (grey dotted) and observed immunity levels at baseline (black dot). The light shaded regions represent the corresponding 95% Bayesian credible intervals.

**Extended Data Figure 10:**
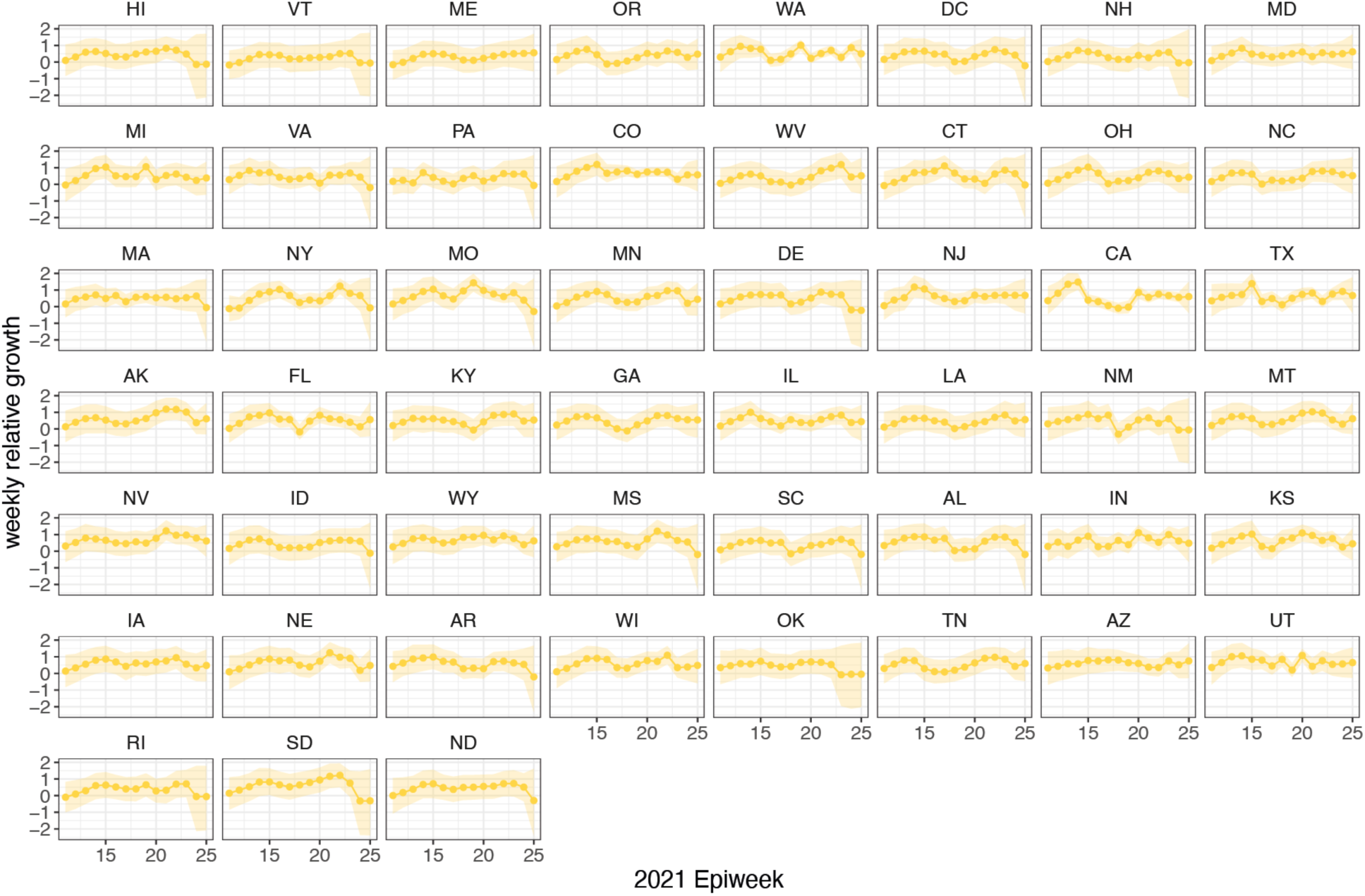
Time-varying relative growth of Delta (on the log odds scale; Methods) for all US states. The light shaded regions represent the corresponding 95% Bayesian credible intervals.

**Extended Data Figure 11:**
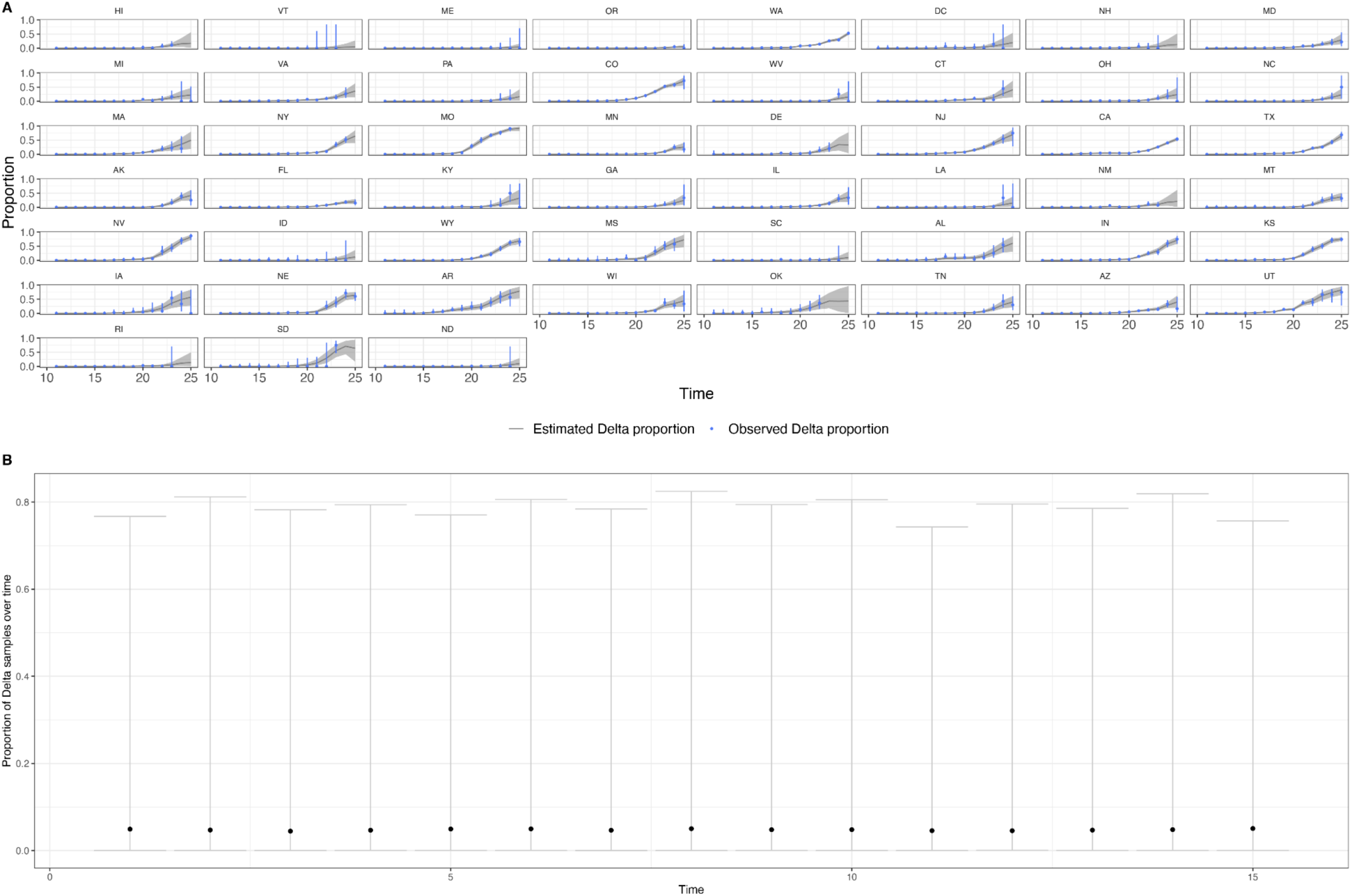
a) Posterior predictive check plotting the observed and predicted proportion of Delta samples for US states. The grey area and blue vertical lines are the 95% Bayesian credible intervals for the observed and predicted proportions. b) Prior predictive check plotting the estimated proportion of Delta samples over time in a single region. The relatively low medians (black dots) indicate the prior assumption of an initially low proportion of Delta samples, and the wide intervals indicate how the priors are relatively uninformative of the Delta proportion.

**Extended Data Figure 12:**
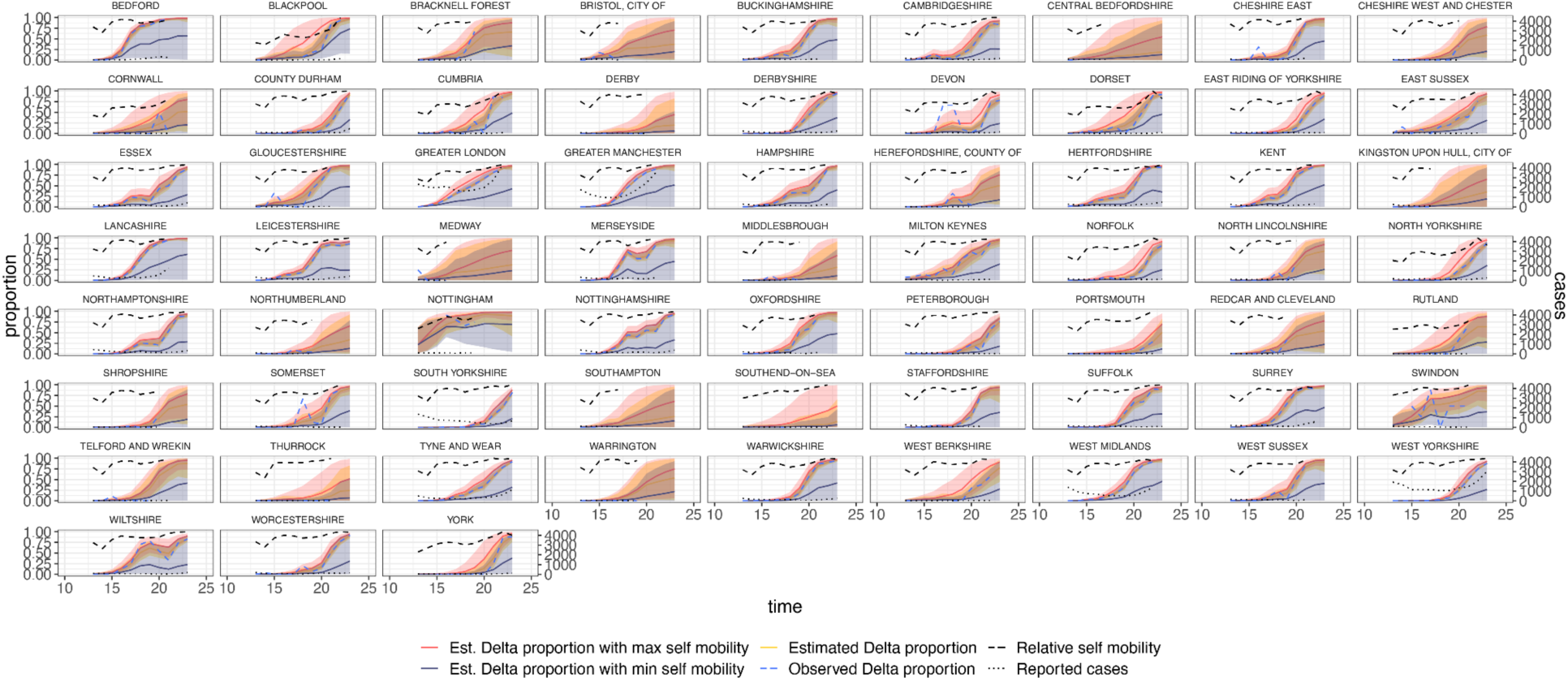
Estimated and observed proportions of Delta variant samples across UTLAs in England (yellow and blue dashed respectively), for various counterfactual scenarios: minimum (purple) and maximum relative self mobility (red), observed relative self-mobility (black dashed), and number of reported cases (black dotted dashed) . The light shaded regions represent the corresponding 95% Bayesian credible intervals.

**Extended Data Figure 13:**
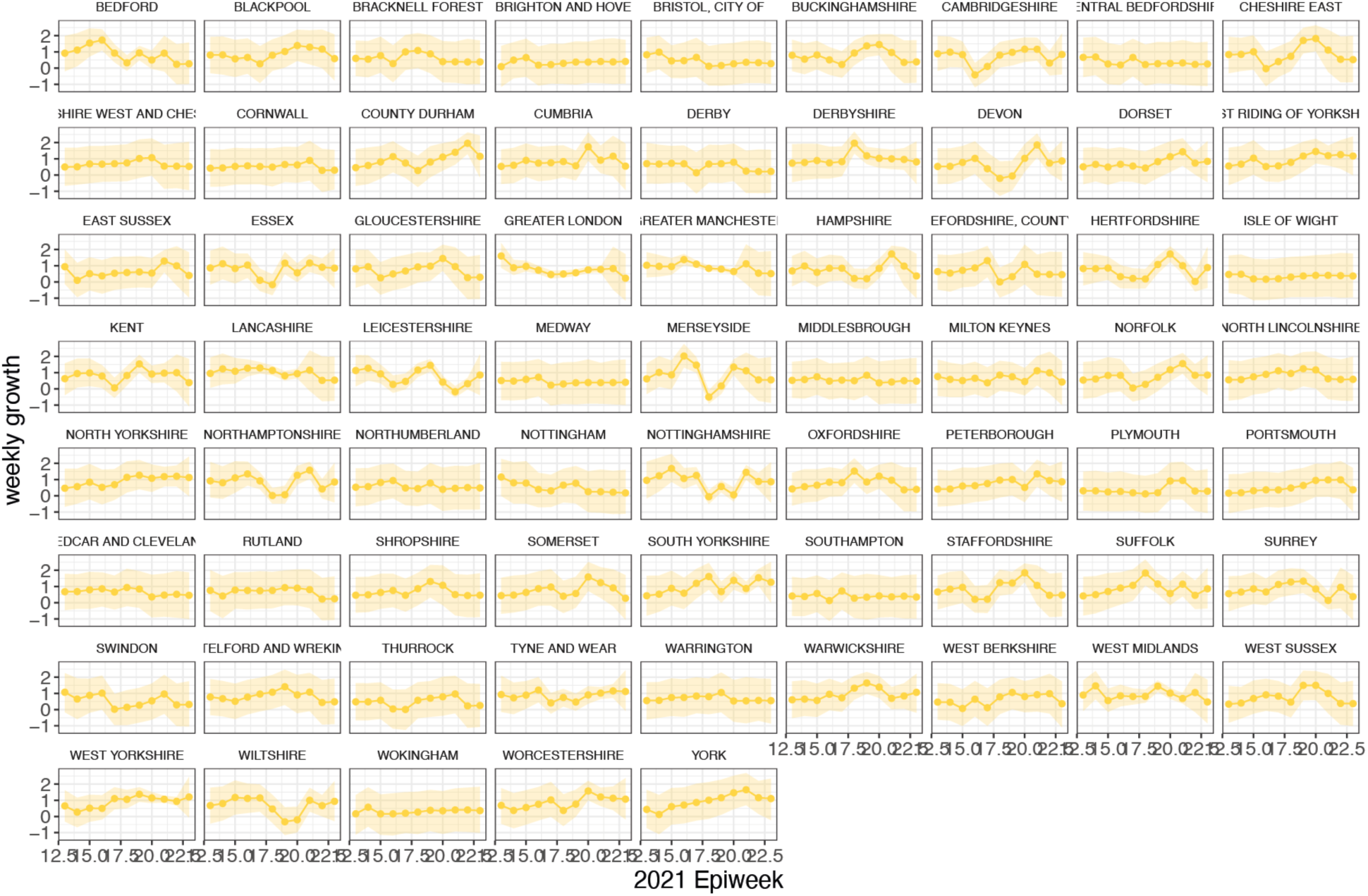
Time-varying relative growth of Delta (on the log odds scale). The light shaded regions represent the corresponding 95% Bayesian credible intervals.

**Extended Data Figure 14:**
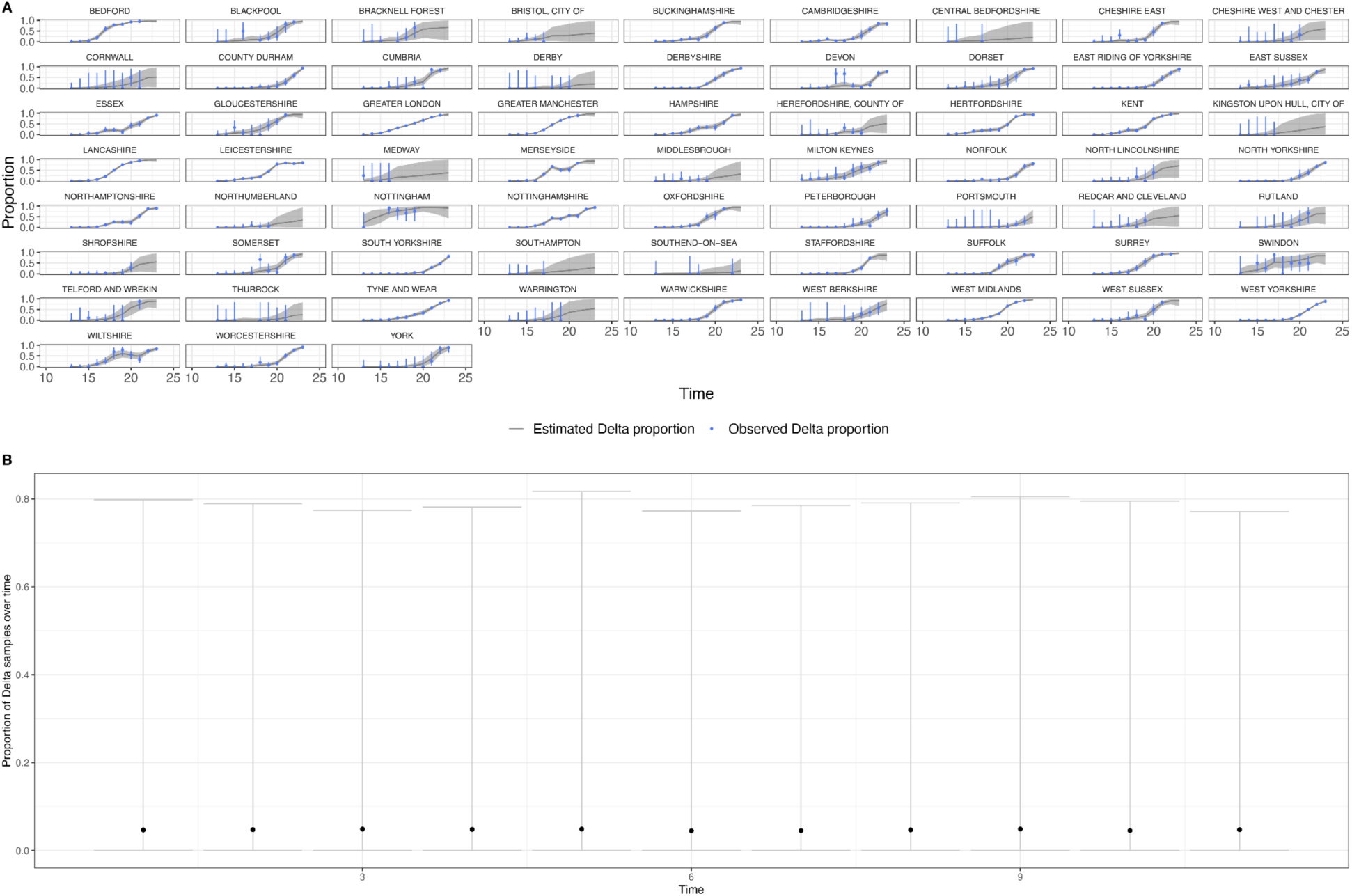
a) Posterior predictive check plotting the observed and predicted proportion of Delta samples for the UTLAs in England. The grey area and blue vertical lines are the 95% Bayesian credible intervals for the observed and predicted proportions. b) Prior predictive check plotting the estimated proportion of Delta samples over time in a single region. The relatively low medians (black dots) indicate the prior assumption of an initially low proportion of starting number of Delta samples, and the wide intervals indicate how the priors are relatively uninformative of the Delta proportion.

**Extended Data Figure 15:**
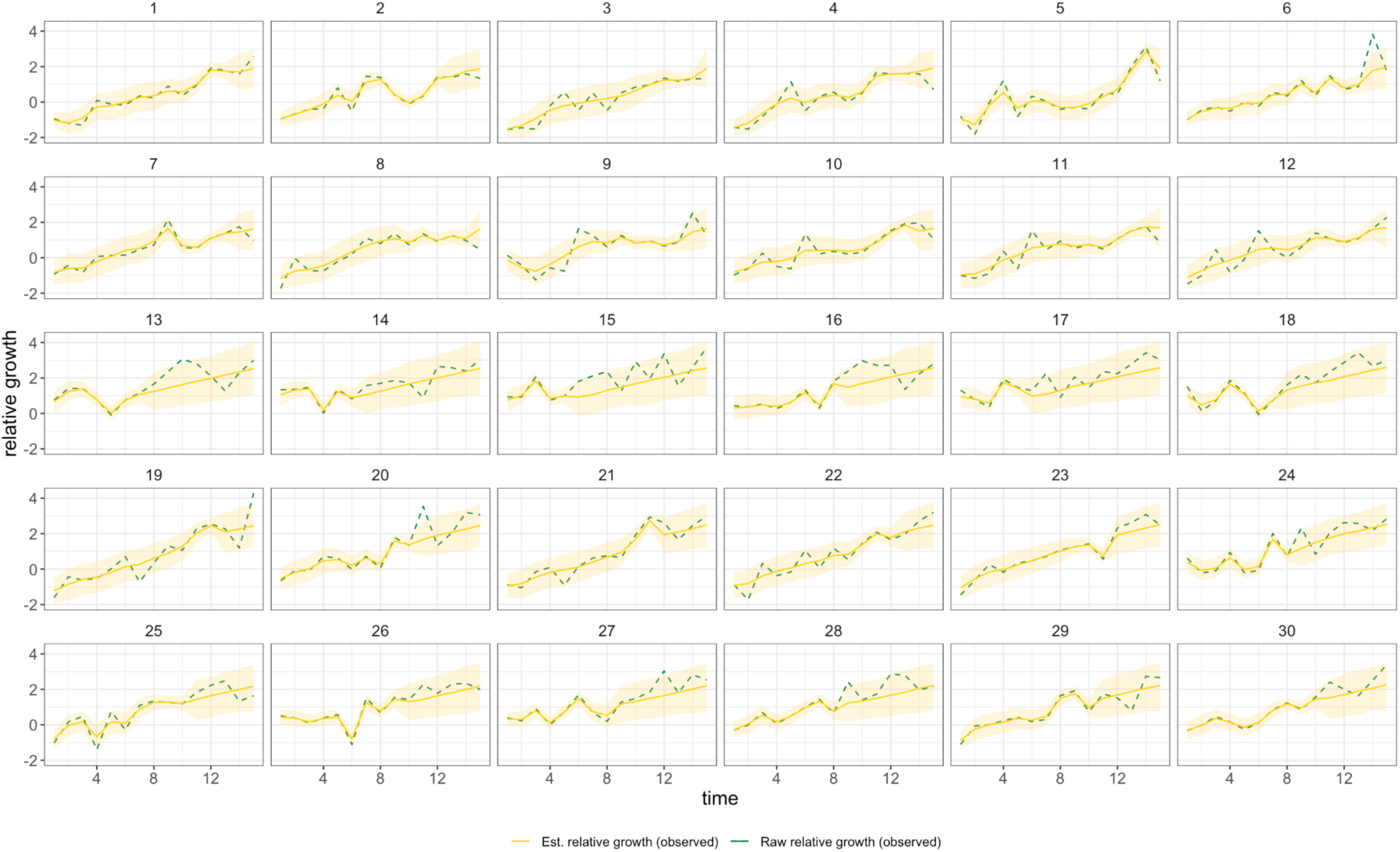
Simulation comparing known vs estimated relative growth rates (see Methods) for hypothetical locations.

## Supplementary Information

**Supplementary Table 1:** The COVID-19 Genomics UK (COG-UK) consortium, June 2021 V.1

**The COVID-19 Genomics UK (COG-UK) consortium June 2021 V.1**

**Funding acquisition, Leadership and supervision, Metadata curation, Project administration, Samples and logistics, Sequencing and analysis, Software and analysis tools, and Visualisation:**

**Samuel C Robson ^13, 84^**

**Funding acquisition, Leadership and supervision, Metadata curation, Project administration, Samples and logistics, Sequencing and analysis, and Software and analysis tools:**

**Thomas R Connor ^11, 74^ and Nicholas J Loman ^43^**

**Leadership and supervision, Metadata curation, Project administration, Samples and logistics, Sequencing and analysis, Software and analysis tools, and Visualisation:**

**Tanya Golubchik ^5^**

**Funding acquisition, Leadership and supervision, Metadata curation, Samples and logistics, Sequencing and analysis, and Visualisation:**

**Rocio T Martinez Nunez ^46^**

**Funding acquisition, Leadership and supervision, Project administration, Samples and logistics, Sequencing and analysis, and Software and analysis tools:**

**David Bonsall ^5^**

**Funding acquisition, Leadership and supervision, Project administration, Sequencing and analysis, Software and analysis tools, and Visualisation:**

**Andrew Rambaut ^104^**

**Funding acquisition, Metadata curation, Project administration, Samples and logistics, Sequencing and analysis, and Software and analysis tools:**

**Luke B Snell ^12^**

**Leadership and supervision, Metadata curation, Project administration, Samples and logistics, Software and analysis tools, and Visualisation:**

**Rich Livett ^116^**

**Funding acquisition, Leadership and supervision, Metadata curation, Project administration, and Samples and logistics:**

**Catherine Ludden ^20, 70^**

**Funding acquisition, Leadership and supervision, Metadata curation, Samples and logistics, and Sequencing and analysis:**

**Sally Corden ^74^ and Eleni Nastouli ^96, 95, 30^**

**Funding acquisition, Leadership and supervision, Metadata curation, Sequencing and analysis, and Software and analysis tools:**

**Gaia Nebbia ^12^**

**Funding acquisition, Leadership and supervision, Project administration, Samples and logistics, and Sequencing and analysis:**

**Ian Johnston ^116^**

**Leadership and supervision, Metadata curation, Project administration, Samples and logistics, and Sequencing and analysis:**

**Katrina Lythgoe ^5^, M. Estee Torok ^19, 20^ and Ian G Goodfellow ^24^**

**Leadership and supervision, Metadata curation, Project administration, Samples and logistics, and Visualisation:**

**Jacqui A Prieto ^97, 82^ and Kordo Saeed ^97, 83^**

**Leadership and supervision, Metadata curation, Project administration, Sequencing and analysis, and Software and analysis tools:**

**David K Jackson ^116^**

**Leadership and supervision, Metadata curation, Samples and logistics, Sequencing and analysis, and Visualisation:**

**Catherine Houlihan ^96, 94^**

**Leadership and supervision, Metadata curation, Sequencing and analysis, Software and analysis tools, and Visualisation:**

**Dan Frampton ^94, 95^**

**Metadata curation, Project administration, Samples and logistics, Sequencing and analysis, and Software and analysis tools:**

**William L Hamilton ^19^ and Adam A Witney ^41^**

**Funding acquisition, Samples and logistics, Sequencing and analysis, and Visualisation:**

**Giselda Bucca ^101^**

**Funding acquisition, Leadership and supervision, Metadata curation, and Project administration:**

**Cassie F Pope ^40, 41^**

**Funding acquisition, Leadership and supervision, Metadata curation, and Samples and logistics:**

**Catherine Moore ^74^**

**Funding acquisition, Leadership and supervision, Metadata curation, and Sequencing and analysis:**

**Emma C Thomson ^53^**

**Funding acquisition, Leadership and supervision, Project administration, and Samples and logistics:**

**Ewan M Harrison ^116, 102^**

**Funding acquisition, Leadership and supervision, Sequencing and analysis, and Visualisation:**

**Colin P Smith ^101^**

**Leadership and supervision, Metadata curation, Project administration, and Sequencing and analysis:**

**Fiona Rogan ^77^**

**Leadership and supervision, Metadata curation, Project administration, and Samples and logistics:**

**Shaun M Beckwith ^6^, Abigail Murray ^6^, Dawn Singleton ^6^, Kirstine Eastick ^37^, Liz A Sheridan ^98^, Paul Randell ^99^, Leigh M Jackson ^105^, Cristina V Ariani ^116^ and Sónia Gonçalves ^116^**

**Leadership and supervision, Metadata curation, Samples and logistics, and Sequencing and analysis:**

**Derek J Fairley ^3, 77^, Matthew W Loose ^18^ and Joanne Watkins ^74^**

**Leadership and supervision, Metadata curation, Samples and logistics, and Visualisation:**

**Samuel Moses ^25, 106^**

**Leadership and supervision, Metadata curation, Sequencing and analysis, and Software and analysis tools:**

**Sam Nicholls ^43^, Matthew Bull ^74^ and Roberto Amato ^116^**

**Leadership and supervision, Project administration, Samples and logistics, and Sequencing and analysis:**

**Darren L Smith ^36, 65, 66^**

**Leadership and supervision, Sequencing and analysis, Software and analysis tools, and Visualisation:**

**David M Aanensen ^14, 116^ and Jeffrey C Barrett ^116^**

**Metadata curation, Project administration, Samples and logistics, and Sequencing and analysis:**

**Dinesh Aggarwal ^20, 116, 70^, James G Shepherd ^53^, Martin D Curran ^71^ and Surendra Parmar ^71^**

**Metadata curation, Project administration, Sequencing and analysis, and Software and analysis tools:**

**Matthew D Parker ^109^**

**Metadata curation, Samples and logistics, Sequencing and analysis, and Software and analysis tools:**

**Catryn Williams ^74^**

**Metadata curation, Samples and logistics, Sequencing and analysis, and Visualisation:**

**Sharon Glaysher ^68^**

**Metadata curation, Sequencing and analysis, Software and analysis tools, and Visualisation:**

**Anthony P Underwood ^14, 116^, Matthew Bashton ^36, 65^, Nicole Pacchiarini ^74^, Katie F Loveson ^84^ and Matthew Byott ^95, 96^**

**Project administration, Sequencing and analysis, Software and analysis tools, and Visualisation:**

**Alessandro M Carabelli ^20^**

**Funding acquisition, Leadership and supervision, and Metadata curation:**

**Kate E Templeton ^56, 104^**

**Funding acquisition, Leadership and supervision, and Project administration:**

**Thushan I de Silva ^109^, Dennis Wang ^109^, Cordelia F Langford ^116^ and John Sillitoe ^116^**

**Funding acquisition, Leadership and supervision, and Samples and logistics:**

**Rory N Gunson ^55^**

**Funding acquisition, Leadership and supervision, and Sequencing and analysis:**

**Simon Cottrell ^74^, Justin O’Grady ^75, 103^ and Dominic Kwiatkowski ^116, 108^**

**Leadership and supervision, Metadata curation, and Project administration:**

**Patrick J Lillie ^37^**

**Leadership and supervision, Metadata curation, and Samples and logistics:**

**Nicholas Cortes ^33^, Nathan Moore ^33^, Claire Thomas ^33^, Phillipa J Burns ^37^, Tabitha W Mahungu ^80^ and Steven Liggett ^86^**

**Leadership and supervision, Metadata curation, and Sequencing and analysis:**

**Angela H Beckett ^13, 81^ and Matthew TG Holden ^73^**

**Leadership and supervision, Project administration, and Samples and logistics:**

**Lisa J Levett ^34^, Husam Osman ^70, 35^ and Mohammed O Hassan-Ibrahim ^99^**

**Leadership and supervision, Project administration, and Sequencing and analysis:**

**David A Simpson ^77^**

**Leadership and supervision, Samples and logistics, and Sequencing and analysis:**

**Meera Chand ^72^, Ravi K Gupta ^102^, Alistair C Darby ^107^ and Steve Paterson ^107^**

**Leadership and supervision, Sequencing and analysis, and Software and analysis tools:**

**Oliver G Pybus ^23^, Erik M Volz ^39^, Daniela de Angelis ^52^, David L Robertson ^53^, Andrew J Page ^75^ and Inigo Martincorena ^116^**

**Leadership and supervision, Sequencing and analysis, and Visualisation:**

**Louise Aigrain ^116^ and Andrew R Bassett ^116^**

**Metadata curation, Project administration, and Samples and logistics:**

**Nick Wong ^50^, Yusri Taha ^89^, Michelle J Erkiert ^99^ and Michael H Spencer Chapman ^116, 102^**

**Metadata curation, Project administration, and Sequencing and analysis:**

**Rebecca Dewar ^56^ and Martin P McHugh ^56, 111^**

**Metadata curation, Project administration, and Software and analysis tools:**

**Siddharth Mookerjee ^38, 57^**

**Metadata curation, Project administration, and Visualisation:**

**Stephen Aplin ^97^, Matthew Harvey ^97^, Thea Sass ^97^, Helen Umpleby ^97^ and Helen Wheeler ^97^**

**Metadata curation, Samples and logistics, and Sequencing and analysis:**

**James P McKenna ^3^, Ben Warne ^9^, Joshua F Taylor ^22^, Yasmin Chaudhry ^24^, Rhys Izuagbe ^24^, Aminu S Jahun ^24^, Gregory R Young ^36, 65^, Claire McMurray ^43^, Clare M McCann ^65, 66^, Andrew Nelson ^65, 66^ and Scott Elliott ^68^**

**Metadata curation, Samples and logistics, and Visualisation:**

**Hannah Lowe ^25^**

**Metadata curation, Sequencing and analysis, and Software and analysis tools:**

**Anna Price ^11^, Matthew R Crown ^65^, Sara Rey ^74^, Sunando Roy ^96^ and Ben Temperton ^105^**

**Metadata curation, Sequencing and analysis, and Visualisation:**

**Sharif Shaaban ^73^ and Andrew R Hesketh ^101^**

**Project administration, Samples and logistics, and Sequencing and analysis:**

**Kenneth G Laing ^41^, Irene M Monahan ^41^ and Judith Heaney ^95, 96, 34^**

**Project administration, Samples and logistics, and Visualisation:**

**Emanuela Pelosi ^97^, Siona Silviera ^97^ and Eleri Wilson-Davies ^97^**

**Samples and logistics, Software and analysis tools, and Visualisation:**

**Helen Fryer ^5^**

**Sequencing and analysis, Software and analysis tools, and Visualization:**

**Helen Adams ^4^, Louis du Plessis ^23^, Rob Johnson ^39^, William T Harvey ^53, 42^, Joseph Hughes ^53^, Richard J Orton ^53^, Lewis G Spurgin ^59^, Yann Bourgeois ^81^, Chris Ruis ^102^, Áine O’Toole ^104^, Marina Gourtovaia ^116^ and Theo Sanderson ^116^**

**Funding acquisition, and Leadership and supervision:**

**Christophe Fraser ^5^, Jonathan Edgeworth ^12^, Judith Breuer ^96, 29^, Stephen L Michell ^105^ and John A Todd ^115^**

**Funding acquisition, and Project administration:**

**Michaela John ^10^ and David Buck ^115^**

**Leadership and supervision, and Metadata curation:**

**Kavitha Gajee ^37^ and Gemma L Kay ^75^**

**Leadership and supervision, and Project administration:**

**Sharon J Peacock ^20, 70^ and David Heyburn ^74^**

**Leadership and supervision, and Samples and logistics:**

**Katie Kitchman ^37^, Alan McNally ^43, 93^, David T Pritchard ^50^, Samir Dervisevic ^58^, Peter Muir ^70^, Esther Robinson ^70, 35^, Barry B Vipond ^70^, Newara A Ramadan ^78^, Christopher Jeanes ^90^, Danni Weldon ^116^, Jana Catalan ^118^ and Neil Jones ^118^**

**Leadership and supervision, and Sequencing and analysis:**

**Ana da Silva Filipe ^53^, Chris Williams ^74^, Marc Fuchs ^77^, Julia Miskelly ^77^, Aaron R Jeffries ^105^, Karen Oliver ^116^ and Naomi R Park ^116^**

**Metadata curation, and Samples and logistics:**

**Amy Ash ^1^, Cherian Koshy ^1^, Magdalena Barrow ^7^, Sarah L Buchan ^7^, Anna Mantzouratou ^7^, Gemma Clark ^15^, Christopher W Holmes ^16^, Sharon Campbell ^1^**^7^**, Thomas Davis ^21^, Ngee Keong Tan ^22^, Julianne R Brown ^29^, Kathryn A Harris ^29, 2^, Stephen P Kidd ^33^, Paul R Grant ^34^, Li Xu-McCrae ^35^, Alison Cox ^38, 63^, Pinglawathee Madona ^38, 63^, Marcus Pond ^38, 63^, Paul A Randell ^38, 63^, Karen T Withell ^48^, Cheryl Williams ^51^, Clive Graham ^60^, Rebecca Denton-Smith ^62^, Emma Swindells ^62^, Robyn Turnbull ^62^, Tim J Sloan ^67^, Andrew Bosworth ^70, 35^, Stephanie Hutchings ^70^, Hannah M Pymont ^70^, Anna Casey ^76^, Liz Ratcliffe ^76^, Christopher R Jones ^79, 105^, Bridget A Knight ^79, 105^, Tanzina Haque ^80^, Jennifer Hart ^80^, Dianne Irish-Tavares ^80^, Eric Witele ^80^, Craig Mower ^86^, Louisa K Watson ^86^, Jennifer Collins ^89^, Gary Eltringham ^89^, Dorian Crudgington ^98^, Ben Macklin ^98^, Miren Iturriza-Gomara ^107^, Anita O Lucaci ^107^ and Patrick C McClure ^113^**

**Metadata curation, and Sequencing and analysis:**

**Matthew Carlile ^18^, Nadine Holmes ^18^, Christopher Moore ^18^, Nathaniel Storey ^29^, Stefan Rooke ^73^, Gonzalo Yebra ^73^, Noel Craine ^74^, Malorie Perry ^74^, Nabil-Fareed Alikhan ^75^, Stephen Bridgett ^77^, Kate F Cook ^84^, Christopher Fearn ^84^, Salman Goudarzi ^84^, Ronan A Lyons ^88^, Thomas Williams ^104^, Sam T Haldenby ^107^, Jillian Durham ^116^ and Steven Leonard ^116^**

**Metadata curation, and Software and analysis tools:**

**Robert M Davies ^116^**

**Project administration, and Samples and logistics:**

**Rahul Batra ^12^, Beth Blane ^20^, Moira J Spyer ^30, 95, 96^, Perminder Smith ^32, 112^, Mehmet Yavus ^85, 109^, Rachel J Williams ^96^, Adhyana IK Mahanama ^97^, Buddhini Samaraweera ^97^, Sophia T Girgis ^102^, Samantha E Hansford ^109^, Angie Green ^115^, Charlotte Beaver ^116^, Katherine L Bellis ^116, 102^, Matthew J Dorman ^116^, Sally Kay ^116^, Liam Prestwood ^116^ and Shavanthi Rajatileka ^116^**

**Project administration, and Sequencing and analysis:**

**Joshua Quick ^43^**

**Project administration, and Software and analysis tools:**

**Radoslaw Poplawski ^43^**

**Samples and logistics, and Sequencing and analysis:**

**Nicola Reynolds ^8^, Andrew Mack ^11^, Arthur Morriss ^11^, Thomas Whalley ^11^, Bindi Patel ^12^, Iliana Georgana ^24^, Myra Hosmillo ^24^, Malte L Pinckert ^24^, Joanne Stockton ^43^, John H Henderson ^65^, Amy Hollis ^65^, William Stanley ^65^, Wen C Yew ^65^, Richard Myers ^72^, Alicia Thornton ^72^, Alexander Adams ^74^, Tara Annett ^74^, Hibo Asad ^74^, Alec Birchley ^74^, Jason Coombes ^74^, Johnathan M Evans ^74^, Laia Fina ^74^, Bree Gatica-Wilcox ^74^, Lauren Gilbert ^74^, Lee Graham ^74^, Jessica Hey ^74^, Ember Hilvers ^74^, Sophie Jones ^74^, Hannah Jones ^74^, Sara Kumziene-Summerhayes ^74^, Caoimhe McKerr ^74^, Jessica Powell ^74^, Georgia Pugh ^74^, Sarah Taylor ^74^, Alexander J Trotter ^75^, Charlotte A Williams ^96^, Leanne M Kermack ^102^, Benjamin H Foulkes ^109^, Marta Gallis ^109^, Hailey R Hornsby ^109^, Stavroula F Louka ^109^, Manoj Pohare ^109^, Paige Wolverson ^109^, Peijun Zhang ^109^, George MacIntyre-Cockett ^115^, Amy Trebes ^115^, Robin J Moll ^116^, Lynne Ferguson ^117^, Emily J Goldstein ^117^, Alasdair Maclean ^117^ and Rachael Tomb ^117^**

**Samples and logistics, and Software and analysis tools:**

**Igor Starinskij ^53^**

**Sequencing and analysis, and Software and analysis tools:**

**Laura Thomson ^5^, Joel Southgate ^11, 74^, Moritz UG Kraemer ^23^, Jayna Raghwani ^23^, Alex E Zarebski ^23^, Olivia Boyd ^39^, Lily Geidelberg ^39^, Chris J Illingworth ^52^, Chris Jackson ^52^, David Pascall ^52^, Sreenu Vattipally ^53^, Timothy M Freeman ^109^, Sharon N Hsu ^109^, Benjamin B Lindsey ^109^, Keith James ^116^, Kevin Lewis ^116^, Gerry Tonkin-Hill ^116^ and Jaime M Tovar-Corona ^116^**

**Sequencing and analysis, and Visualisation:**

**MacGregor Cox ^20^**

**Software and analysis tools, and Visualisation:**

**Khalil Abudahab ^14, 116^, Mirko Menegazzo ^14^, Ben EW Taylor MEng ^14, 116^, Corin A Yeats ^14^, Afrida Mukaddas ^53^, Derek W Wright ^53^, Leonardo de Oliveira Martins ^75^, Rachel Colquhoun ^104^, Verity Hill ^104^, Ben Jackson ^104^, JT McCrone ^104^, Nathan Medd ^104^, Emily Scher ^104^ and Jon-Paul Keatley ^116^**

**Leadership and supervision:**

**Tanya Curran ^3^, Sian Morgan ^10^, Patrick Maxwell ^20^, Ken Smith ^20^, Sahar Eldirdiri ^21^, Anita Kenyon ^21^, Alison H Holmes ^38, 57^, James R Price ^38, 57^, Tim Wyatt ^69^, Alison E Mather ^75^, Timofey Skvortsov ^77^ and John A Hartley ^96^**

**Metadata curation:**

**Martyn Guest ^11^, Christine Kitchen ^11^, Ian Merrick ^11^, Robert Munn ^11^, Beatrice Bertolusso ^33^, Jessica Lynch ^33^, Gabrielle Vernet ^33^, Stuart Kirk ^34^, Elizabeth Wastnedge ^56^, Rachael Stanley ^58^, Giles Idle ^64^, Declan T Bradley ^69, 77^, Jennifer Poyner ^79^ and Matilde Mori ^110^**

**Project administration:**

**Owen Jones ^11^, Victoria Wright ^18^, Ellena Brooks ^20^, Carol M Churcher ^20^, Mireille Fragakis ^20^, Katerina Galai ^20, 70^, Andrew Jermy ^20^, Sarah Judges ^20^, Georgina M McManus ^20^, Kim S Smith ^20^, Elaine Westwick ^20^, Stephen W Attwood ^23^, Frances Bolt ^38, 57^, Alisha Davies ^74^, Elen De Lacy ^74^, Fatima Downing ^74^, Sue Edwards ^74^, Lizzie Meadows ^75^, Sarah Jeremiah ^97^, Nikki Smith ^109^ and Luke Foulser ^116^**

**Samples and logistics:**

**Themoula Charalampous ^12, 46^, Amita Patel ^12^, Louise Berry ^15^, Tim Boswell ^15^, Vicki M Fleming ^15^, Hannah C Howson-Wells ^15^, Amelia Joseph ^15^, Manjinder Khakh ^15^, Michelle M Lister ^15^, Paul W Bird ^16^, Karlie Fallon ^16^, Thomas Helmer ^16^, Claire L McMurray ^16^, Mina Odedra ^16^, Jessica Shaw ^16^, Julian W Tang ^16^, Nicholas J Willford ^16^, Victoria Blakey ^17^, Veena Raviprakash ^17^, Nicola Sheriff ^17^, Lesley-Anne Williams** ^17^**, Theresa Feltwell ^20^, Luke Bedford ^26^, James S Cargill ^27^, Warwick Hughes ^27^, Jonathan Moore ^28^, Susanne Stonehouse ^28^, Laura Atkinson** ^29^**, Jack CD Lee** ^29^**, Dr Divya Shah ^29^, Adela Alcolea-Medina ^32, 112^, Natasha Ohemeng-Kumi ^32, 112^, John Ramble ^32, 1^**^12^**, Jasveen Sehmi ^32, 112^, Rebecca Williams ^33^, Wendy Chatterton ^34^, Monika Pusok ^34^, William Everson ^37^, Anibolina Castigador ^44^, Emily Macnaughton ^44^, Kate El Bouzidi ^45^, Temi Lampejo ^45^, Malur Sudhanva ^45^, Cassie Breen ^47^, Graciela Sluga ^48^, Shazaad SY Ahmad ^49, 70^, Ryan P George ^49^, Nicholas W Machin ^49, 70^, Debbie Binns ^50^, Victoria James ^50^, Rachel Blacow ^55^, Lindsay Coupland ^58^, Louise Smith ^59^, Edward Barton ^60^, Debra Padgett ^60^, Garren Scott ^60^, Aidan Cross ^61^, Mariyam Mirfenderesky ^61^, Jane Greenaway ^62^, Kevin Cole ^64^, Phillip Clarke ^67^, Nichola Duckworth ^67^, Sarah Walsh ^67^, Kelly Bicknell ^68^, Robert Impey ^68^, Sarah Wyllie ^68^, Richard Hopes ^70^, Chloe Bishop ^72^, Vicki Chalker ^72^, Ian Harrison ^72^, Laura Gifford ^74^, Zoltan Molnar ^77^, Cressida Auckland ^79^, Cariad Evans ^85, 109^, Kate Johnson ^85, 109^, David G Partridge ^85, 109^, Mohammad Raza ^85, 109^, Paul Baker ^86^, Stephen Bonner ^86^, Sarah Essex ^86^, Leanne J Murray ^86^, Andrew I Lawton ^87^, Shirelle Burton-Fanning ^89^, Brendan AI Payne ^89^, Sheila Waugh ^89^, Andrea N Gomes ^91^, Maimuna Kimuli ^91^, Darren R Murray ^91^, Paula Ashfield ^92^, Donald Dobie ^92^, Fiona Ashford ^93^, Angus Best ^93^, Liam Crawford ^93^, Nicola Cumley ^93^, Megan Mayhew ^93^, Oliver Megram ^93^, Jeremy Mirza ^93^, Emma Moles-Garcia ^93^, Benita Percival ^93^, Megan Driscoll ^96^, Leah Ensell ^96^, Helen L Lowe ^96^, Laurentiu Maftei ^96^, Matteo Mondani ^96^, Nicola J Chaloner ^99^, Benjamin J Cogger ^99^, Lisa J Easton ^99^, Hannah Huckson ^99^, Jonathan Lewis ^99^, Sarah Lowdon ^99^, Cassandra S Malone ^99^, Florence Munemo ^99^, Manasa Mutingwende ^99^, Roberto Nicodemi ^99^, Olga Podplomyk ^99^, Thomas Somassa ^99^, Andrew Beggs ^100^, Alex Richter ^100^, Claire Cormie ^102^, Joana Dias ^102^, Sally Forrest ^102^, Ellen E Higginson ^102^, Mailis Maes ^102^, Jamie Young ^102^, Rose K Davidson ^103^, Kathryn A Jackson ^107^, Lance Turtle ^107^, Alexander J Keeley ^109^, Jonathan Ball ^113^, Timothy Byaruhanga ^113^, Joseph G Chappell ^113^, Jayasree Dey ^113^, Jack D Hill ^113^, Emily J Park ^113^, Arezou Fanaie ^114^, Rachel A Hilson ^114^, Geraldine Yaze ^114^ and Stephanie Lo ^116^**

**Sequencing and analysis:**

**Safiah Afifi ^10^, Robert Beer ^10^, Joshua Maksimovic ^10^, Kathryn McCluggage ^10^, Karla Spellman ^10^, Catherine Bresner ^11^, William Fuller ^11^, Angela Marchbank ^11^, Trudy Workman ^11^, Ekaterina Shelest ^13, 81^, Johnny Debebe ^18^, Fei Sang ^18^, Marina Escalera Zamudio ^23^, Sarah Francois ^23^, Bernardo Gutierrez ^23^, Tetyana I Vasylyeva ^23^, Flavia Flaviani ^31^, Manon Ragonnet-Cronin ^39^, Katherine L Smollett ^42^, Alice Broos ^53^, Daniel Mair ^53^, Jenna Nichols ^53^, Kyriaki Nomikou ^53^, Lily Tong ^53^, Ioulia Tsatsani ^53^, Sarah O’Brien ^54^, Steven Rushton ^54^, Roy Sanderson ^54^, Jon Perkins ^55^, Seb Cotton ^56^, Abbie Gallagher ^56^, Elias Allara ^70, 102^, Clare Pearson ^70, 102^, David Bibby ^72^, Gavin Dabrera ^72^, Nicholas Ellaby ^72^, Eileen Gallagher ^72^, Jonathan Hubb ^72^, Angie Lackenby ^72^, David Lee ^72^, Nikos Manesis ^72^, Tamyo Mbisa ^72^, Steven Platt ^72^, Katherine A Twohig ^72^, Mari Morgan ^74^, Alp Aydin ^75^, David J Baker ^75^, Ebenezer Foster-Nyarko ^75^, Sophie J Prosolek ^75^, Steven Rudder ^75^, Chris Baxter ^77^, Sílvia F Carvalho ^77^, Deborah Lavin ^77^, Arun Mariappan ^77^, Clara Radulescu ^77^, Aditi Singh ^77^, Miao Tang ^77^, Helen Morcrette ^79^, Nadua Bayzid ^96^, Marius Cotic ^96^, Carlos E Balcazar ^104^, Michael D Gallagher ^104^, Daniel Maloney ^104^, Thomas D Stanton ^104^, Kathleen A Williamson ^104^, Robin Manley ^105^, Michelle L Michelsen ^105^, Christine M Sambles ^105^, David J Studholme ^105^, Joanna Warwick-Dugdale ^105^, Richard Eccles ^107^, Matthew Gemmell ^107^, Richard Gregory ^107^, Margaret Hughes ^107^, Charlotte Nelson ^107^, Lucille Rainbow ^107^, Edith E Vamos ^107^, Hermione J Webster ^107^, Mark Whitehead ^107^, Claudia Wierzbicki ^107^, Adrienn Angyal ^109^, Luke R Green ^109^, Max Whiteley ^109^, Emma Betteridge ^116^, Iraad F Bronner ^116^, Ben W Farr ^116^, Scott Goodwin ^116^, Stefanie V Lensing ^116^, Shane A McCarthy ^116, 102^, Michael A Quail ^116^, Diana Rajan ^116^, Nicholas M Redshaw ^116^, Carol Scott ^116^, Lesley Shirley ^116^ and Scott AJ Thurston ^116^**

**Software and analysis tools:**

**Will Rowe ^43^, Amy Gaskin ^74^, Thanh Le-Viet ^75^, James Bonfield ^116^, Jennifier Liddle ^116^ and Andrew Whitwham ^116^**

**1 Barking, Havering and Redbridge University Hospitals NHS Trust, 2 Barts Health NHS Trust, 3 Belfast Health & Social Care Trust, 4 Betsi Cadwaladr University Health Board, 5 Big Data Institute, Nuffield Department of Medicine, University of Oxford, 6 Blackpool Teaching Hospitals NHS Foundation Trust, 7 Bournemouth University, 8 Cambridge Stem Cell Institute, University of Cambridge, 9 Cambridge University Hospitals NHS Foundation Trust, 10 Cardiff and Vale University Health Board, 11 Cardiff University, 12 Centre for Clinical Infection and Diagnostics Research, Department of Infectious Diseases, Guy’s and St Thomas’ NHS Foundation Trust, 13 Centre for Enzyme Innovation, University of Portsmouth, 14 Centre for Genomic Pathogen Surveillance, University of Oxford, 15 Clinical Microbiology Department, Queens Medical Centre, Nottingham University Hospitals NHS Trust, 16 Clinical Microbiology, University Hospitals of Leicester NHS Trust, 17 County Durham and Darlington NHS Foundation Trust, 18 Deep Seq, School of Life Sciences, Queens Medical Centre, University of Nottingham, 19 Department of Infectious Diseases and Microbiology, Cambridge University Hospitals NHS Foundation Trust, 20 Department of Medicine, University of Cambridge, 21 Department of Microbiology, Kettering General Hospital, 22 Department of Microbiology, South West London Pathology, 23 Department of Zoology, University of Oxford, 24 Division of Virology, Department of Pathology, University of Cambridge, 25 East Kent Hospitals University NHS Foundation Trust, 26 East Suffolk and North Essex NHS Foundation Trust, 27 East Sussex Healthcare NHS Trust, 28 Gateshead Health NHS Foundation Trust, 29 Great Ormond Street Hospital for Children NHS Foundation Trust, 30 Great Ormond Street Institute of Child Health (GOS ICH), University College London (UCL), 31 Guy’s and St. Thomas’ Biomedical Research Centre, 32 Guy’s and St. Thomas’ NHS Foundation Trust, 33 Hampshire Hospitals NHS Foundation Trust, 34 Health Services Laboratories, 35 Heartlands Hospital, Birmingham, 36 Hub for Biotechnology in the Built Environment, Northumbria University, 37 Hull University Teaching Hospitals NHS Trust, 38 Imperial College Healthcare NHS Trust, 39 Imperial College London, 40 Infection Care Group, St George’s University Hospitals NHS Foundation Trust, 41 Institute for Infection and Immunity, St George’s University of London, 42 Institute of Biodiversity, Animal Health & Comparative Medicine, 43 Institute of Microbiology and Infection, University of Birmingham, 44 Isle of Wight NHS Trust, 45 King’s College Hospital NHS Foundation Trust, 46 King’s College London, 47 Liverpool Clinical Laboratories, 48 Maidstone and Tunbridge Wells NHS Trust, 49 Manchester University NHS Foundation Trust, 50 Microbiology Department, Buckinghamshire Healthcare NHS Trust, 51 Microbiology, Royal Oldham Hospital, 52 MRC Biostatistics Unit, University of Cambridge, 53 MRC-University of Glasgow Centre for Virus Research, 54 Newcastle University, 55 NHS Greater Glasgow and Clyde, 56 NHS Lothian, 57 NIHR Health Protection Research Unit in HCAI and AMR, Imperial College London, 58 Norfolk and Norwich University Hospitals NHS Foundation Trust, 59 Norfolk County Council, 60 North Cumbria Integrated Care NHS Foundation Trust, 61 North Middlesex University Hospital NHS Trust, 62 North Tees and Hartlepool NHS Foundation Trust, 63 North West London Pathology, 64 Northumbria Healthcare NHS Foundation Trust, 65 Northumbria University, 66 NU-OMICS, Northumbria University, 67 Path Links, Northern Lincolnshire and Goole NHS Foundation Trust, 68 Portsmouth Hospitals University NHS Trust, 69 Public Health Agency, Northern Ireland, 70 Public Health England, 71 Public Health England, Cambridge, 72 Public Health England, Colindale, 73 Public Health Scotland, 74 Public Health Wales, 75 Quadram Institute Bioscience, 76 Queen Elizabeth Hospital, Birmingham, 77 Queen’s University Belfast, 78 Royal Brompton and Harefield Hospitals, 79 Royal Devon and Exeter NHS Foundation Trust, 80 Royal Free London NHS Foundation Trust, 81 School of Biological Sciences, University of Portsmouth, 82 School of Health Sciences, University of Southampton, 83 School of Medicine, University of Southampton, 84 School of Pharmacy & Biomedical Sciences, University of Portsmouth, 85 Sheffield Teaching Hospitals NHS Foundation Trust, 86 South Tees Hospitals NHS Foundation Trust, 87 Southwest Pathology Services, 88 Swansea University, 89 The Newcastle upon Tyne Hospitals NHS Foundation Trust, 90 The Queen Elizabeth Hospital King’s Lynn NHS Foundation Trust, 91 The Royal Marsden NHS Foundation Trust, 92 The Royal Wolverhampton NHS Trust, 93 Turnkey Laboratory, University of Birmingham, 94 University College London Division of Infection and Immunity, 95 University College London Hospital Advanced Pathogen Diagnostics Unit, 96 University College London Hospitals NHS Foundation Trust, 97 University Hospital Southampton NHS Foundation Trust, 98 University Hospitals Dorset NHS Foundation Trust, 99 University Hospitals Sussex NHS Foundation Trust, 100 University of Birmingham, 101 University of Brighton, 102 University of Cambridge, 103 University of East Anglia, 104 University of Edinburgh, 105 University of Exeter, 106 University of Kent, 107 University of Liverpool, 108 University of Oxford, 109 University of Sheffield, 110 University of Southampton, 111 University of St Andrews, 112 Viapath, Guy’s and St Thomas’ NHS Foundation Trust, and King’s College Hospital NHS Foundation Trust, 113 Virology, School of Life Sciences, Queens Medical Centre, University of Nottingham, 114 Watford General Hospital, 115 Wellcome Centre for Human Genetics, Nuffield Department of Medicine, University of Oxford, 116 Wellcome Sanger Institute, 117 West of Scotland Specialist Virology Centre, NHS Greater Glasgow and Clyde, 118 Whittington Health NHS Trust**

